# *Veillonella atypica* supplementation reduces fatigue interference and increases voluntary physical activity: A randomized controlled trial with mechanistic validation in mice

**DOI:** 10.1101/2025.11.03.25339441

**Authors:** Ebru Aras-Hira, Marina Santiago, Natalie Casto, Kayla Weng, Loc-Duyen Pham, Paige Oliver, Marie Crisel Erfe, Carolina Barsa, Noah Craft, Jonathan R. Scheiman, Aleksandar D. Kostic

**Affiliations:** Section on Pathophysiology and Molecular Pharmacology, Joslin Diabetes Center, Boston, MA, USA; Department of Microbiology, Harvard Medical School, Boston, MA, USA; FitBiomics Inc, New York City, NY, USA; People Science Inc, Los Angeles, LA, USA

**Keywords:** *Veillonella atypica*, voluntary exercise, exercise motivation, fatigue, gut-brain axis, microbiome, randomized controlled trial, striatal dopamine

## Abstract

**Background:** The gut microbiome has been implicated in exercise performance and, more recently, in neural pathways that govern exercise motivation. *Veillonella atypica*—a lactate-utilizing bacterium enriched in elite athletes— enhances forced treadmill performance in mice through lactate-to-propionate metabolism, yet effects on voluntary physical activity, the clinically relevant behavior, remain unknown. We tested whether supplementation with *V. atypica* reduces fatigue burden and increases voluntary physical activity in humans, with mechanistic validation in mice.

**Methods:** We conducted a randomized, placebo-controlled, app-mediated trial in healthy adults (8-week protocol: 2-week baseline, 4-week supplementation, 2-week washout; n=151 meeting compliance criteria). Participants received high-dose *V. atypica*, low-dose *V. atypica*, or placebo; dose groups were pooled after confirming similar responses. Primary endpoints were Multidimensional Fatigue Inventory (MFI-20) subscales assessed pre- and post-supplementation; secondary endpoints included weekly surveys (fatigue interference, physical activity hours) and daily surveys (sleep quality, energy). In parallel, we profiled voluntary wheel running in mice randomized to PBS, *V. atypica*, or *Lactobacillus johnsonii* (peanut butter delivery), with striatal dopamine quantification at endpoint.

**Results:** In humans (n=146-149 depending on outcome), weekly longitudinal assessments revealed *V. atypica*-specific benefits not captured by endpoint comparisons. Relative to placebo, *V. atypica* produced a faster decline in fatigue interference (−0.097 days/week; 95% CI −0.175 to −0.018; *p*=0.016) and a faster increase in self-reported physical-activity hours (+0.357 h/week; 95% CI +0.031 to +0.684; *p*=0.032). Model-predicted effects by week 6 corresponded to approximately 0.6 fewer days with fatigue interference and 2 additional activity hours per week if linear trends continued. By contrast, MFI-based fatigue scores improved over time in both arms without between-group differences (treatment×time interactions all *p*>0.40). Daily measures indicated improved sleep quality during washout in *V. atypica* versus placebo (difference-in-differences +0.197 Likert units; *p*=0.0039), with no differences in sleep duration; odds of high-energy days trended higher with *V. atypica* during washout (OR≈2.36; *p*=0.07).

In mice, mixed-effects models revealed a significant group×time interaction for daily running distance (F=13.31; *p*=2.1×10⁻⁴): *V. atypica* maintained running across 12 weeks (slope not significantly different from zero), whereas PBS and *L. johnsonii* declined significantly. During the final 5 weeks (days 49–84), baseline-adjusted mean running distance was approximately 2.86 km/day higher in *V. atypica* versus PBS (*p*=2.2×10⁻⁴). Striatal dopamine concentrations were elevated in *V. atypica*-treated mice relative to both controls (one-way ANOVA *p*=0.05; *V. atypica* vs PBS *p*=0.03, approximately 30% higher).

**Conclusions:** Across complementary human and murine experiments, *V. atypica* was associated with reduced day-to-day fatigue interference and increased voluntary physical activity, with convergent evidence of sustained behavioral engagement in mice. Preliminary findings of elevated striatal dopamine are consistent with emerging evidence that gut microbiome regulates exercise motivation through dopaminergic signaling, though mechanistic causality remains to be established. These data extend *Veillonella* biology beyond performance physiology to motivational domains and support continued investigation of precision microbiome approaches to address barriers to physical activity.

## INTRODUCTION

Regular physical activity is essential for human health and represents a cornerstone of preventive medicine, reducing risk of cardiovascular disease, type 2 diabetes, obesity, certain cancers, and all-cause mortality [1]. Physical activity also enhances cognitive function, improves mental well-being, and modulates the gut microbiome in ways that support metabolic health [2, 3]. Despite these well-established benefits, nearly one-third of the global adult population—approximately 1.8 billion individuals—fail to meet the World Health Organization recommendation of at least 150 minutes of moderate-intensity physical activity per week [4]. The primary barriers to exercise adherence are not physiological capacity but rather motivational factors, including perceived fatigue, low intrinsic motivation, and high subjective effort cost [5–7]. While traditional exercise physiology has focused on musculoskeletal, cardiovascular, and metabolic determinants of performance, emerging evidence suggests that the gut microbiome may represent a previously unrecognized regulator of exercise motivation and fatigue perception.

The gut microbiome comprises trillions of microorganisms that maintain host homeostasis through regulation of metabolism, immune function, and neural communication [8]. The gut-brain axis—a complex, bidirectional communication network encompassing vagal neural signaling, immune modulation, enteroendocrine hormone secretion, and microbial metabolite transmission—has been implicated in diverse behaviors including anxiety, social interaction, and cognition [9]. More recently, this framework has been extended to physical activity. Exercise alters gut microbial composition and metabolic output [2], while gut microbiota in turn influence exercise capacity and performance [3]. Seminal work by Clarke et al. (2014) revealed that elite athletes harbor distinct gut microbiota characterized by increased α-diversity, enrichment of amino acid metabolic pathways, and higher abundance of specific taxa including *Akkermansia muciniphila* and bacteria within the genus *Veillonella* [10]. Subsequent studies confirmed these patterns across diverse athletic populations, with Barton et al. (2018) demonstrating that athletes exhibit elevated fecal short-chain fatty acid (SCFA) concentrations, particularly acetate, propionate, and butyrate—metabolites with known neuroactive properties [11].

The functional significance of *Veillonella* enrichment in athletes was established by our previous work, in which we isolated *Veillonella atypica* from the stool of elite Boston Marathon runners and tested its effects on exercise performance in mice [12]. Oral gavage with *V. atypica* significantly increased treadmill running time to exhaustion compared to a lactic-acid producing bacterial control. This performance enhancement was mechanistically linked to *V. atypica*’s unique metabolic capability: the bacterium specializes in consuming lactate—a byproduct of anaerobic glycolysis that accumulates during intense exercise—and converting it via the succinate (methylmalonyl-CoA) pathway to propionate, a short-chain fatty acid [13]. Cecal propionate concentrations were elevated following *V. atypica* supplementation, establishing a lactate → *V. atypica* → propionate metabolic axis. Propionate itself, when administered exogenously, recapitulated a portion of the performance benefit, supporting its role as a bioactive mediator. This work positioned *V. atypica* as a precision microbiome target—a bacterium with a defined metabolic function relevant to exercise physiology.

A critical limitation of prior microbiome-exercise research, including our own, is the focus on *forced* exercise paradigms—specifically, treadmill running to exhaustion in rodents or laboratory-based maximal exertion tests in humans. These protocols measure physiological capacity under external compulsion and may be limited by peripheral factors such as muscle fatigue, cardiovascular fitness, or metabolic substrate availability [14]. By contrast, *voluntary* physical activity—freely chosen, self-initiated movement—is governed by central neural circuits that integrate motivation, reward valuation, and effort cost computations [15]. The distinction is not merely semantic but has profound clinical implications: the global physical inactivity pandemic is driven primarily by motivational deficits rather than physiological incapacity. Most sedentary individuals are capable of walking for 30 minutes but lack the intrinsic motivation to initiate and sustain the behavior [4]. Voluntary wheel running in rodents—a paradigm in which mice are single-housed with ad libitum access to running wheels and wheel revolutions are logged continuously—provides a model of intrinsic exercise motivation uncoupled from external compulsion [16]. Similarly, self-reported physical activity hours in humans capture volitional engagement rather than forced performance. The neural substrates of voluntary exercise include the striatum, where dopamine signaling regulates both the initiation of goal-directed actions (dorsomedial striatum) and the formation of exercise habits (dorsolateral striatum) [17]. Addressing the physical inactivity crisis requires interventions that enhance motivation to engage in physical activity, not merely capacity to perform when compelled.

A causal role of gut microbiota in exercise motivation was established by Dohnalová et al. (2022) in a landmark study [18]. Germ-free mice and mice treated with broad-spectrum antibiotics exhibited profoundly reduced voluntary wheel running—up to 50% lower than conventionally housed controls—that was rescued by microbial recolonization. Importantly, these effects were specific to voluntary running; forced treadmill performance was unaffected, indicating a motivational rather than physiological performance deficit. Mechanistically, the authors demonstrated that gut microbiota produce fatty acid amides, including N-oleoylethanolamide and N-linoleoylethanolamide, during exercise. These endocannabinoid-like metabolites are elevated in the bloodstream following running and activate TRPV1-expressing spinal sensory neurons. Optogenetic and chemogenetic experiments showed that TRPV1 neuron activity is necessary and sufficient for normal voluntary running levels. These afferent signals project to the nucleus tractus solitarius in the brainstem and ultimately enhance dopamine release in the ventral striatum (nucleus accumbens) during exercise—a neural signature of reward and motivation. Pharmacological blockade of dopamine signaling abolished the motivational effects, establishing striatal dopamine as a critical downstream mediator of microbiome-dependent exercise motivation. This work provided a mechanistic framework linking gut microbial metabolism to defined neural circuits governing behavior.

Despite these advances, three critical gaps remain. First, while our 2019 study demonstrated that *V. atypica* enhances forced treadmill performance in mice through lactate-to-propionate metabolism, whether these effects extend to *voluntary* physical activity—the clinically relevant behavior governed by motivation—is unknown. Second, no human trials have tested whether *V. atypica* supplementation influences physical activity levels, fatigue, or exercise motivation in humans. A recent pilot study by Gross et al. (2024) reported maintained endurance performance in seven individuals receiving *V. atypica* compared to decline in placebo controls, but the sample size was small and did not assess voluntary physical activity engagement [19]. Third, the neural mechanisms by which *V. atypica* might influence exercise behavior remain unexplored. Given that Dohnalová et al. established striatal dopamine as a key mediator of microbiome-dependent exercise motivation, and that propionate—the terminal product of *V. atypica* lactate metabolism—has been shown to cross the blood-brain barrier [20, 21], modulate reward circuitry [22], and protect dopaminergic neurons [23], we hypothesized that *V. atypica* may influence voluntary exercise through a propionate-mediated, dopaminergic mechanism.

Here, we address these gaps through a two-pronged approach combining human translation and mechanistic investigation. We conducted a randomized, placebo-controlled trial in 151 healthy adults to test whether *V. atypica* supplementation reduces fatigue burden and increases voluntary physical activity over an 8-week period, as assessed by validated fatigue inventories, weekly surveys of fatigue interference and physical activity hours, and daily assessments of sleep quality and energy. In parallel, we profiled voluntary wheel running in mice randomized to receive *V. atypica*, PBS, or *Lactobacillus johnsonii* (a common probiotic and lactate-*producing* control bacterium, testing specificity for lactate utilization). At study endpoint, we quantified striatal dopamine concentrations to test the hypothesis that *V. atypica* supplementation modulates the dopaminergic signaling established by Dohnalová et al. to regulate exercise motivation.

## RESULTS

### HUMAN CLINICAL TRIAL OF *VEILLONELLA ATYPICA*

#### Study Design and Participant Flow

We conducted a randomized, placebo-controlled clinical trial to evaluate the effects of the lactate-utilizing bacterium *Veillonella atypica* FB0054 on fatigue, energy, and physical activity in healthy adults. The 8-week protocol comprised a two-week baseline, four-week supplementation, and two-week washout period (**Figure 1**). Participants were recruited through online advertisements and screened for eligibility using an electronic informed consent and survey platform (CHLOE app).

**Figure 1.**
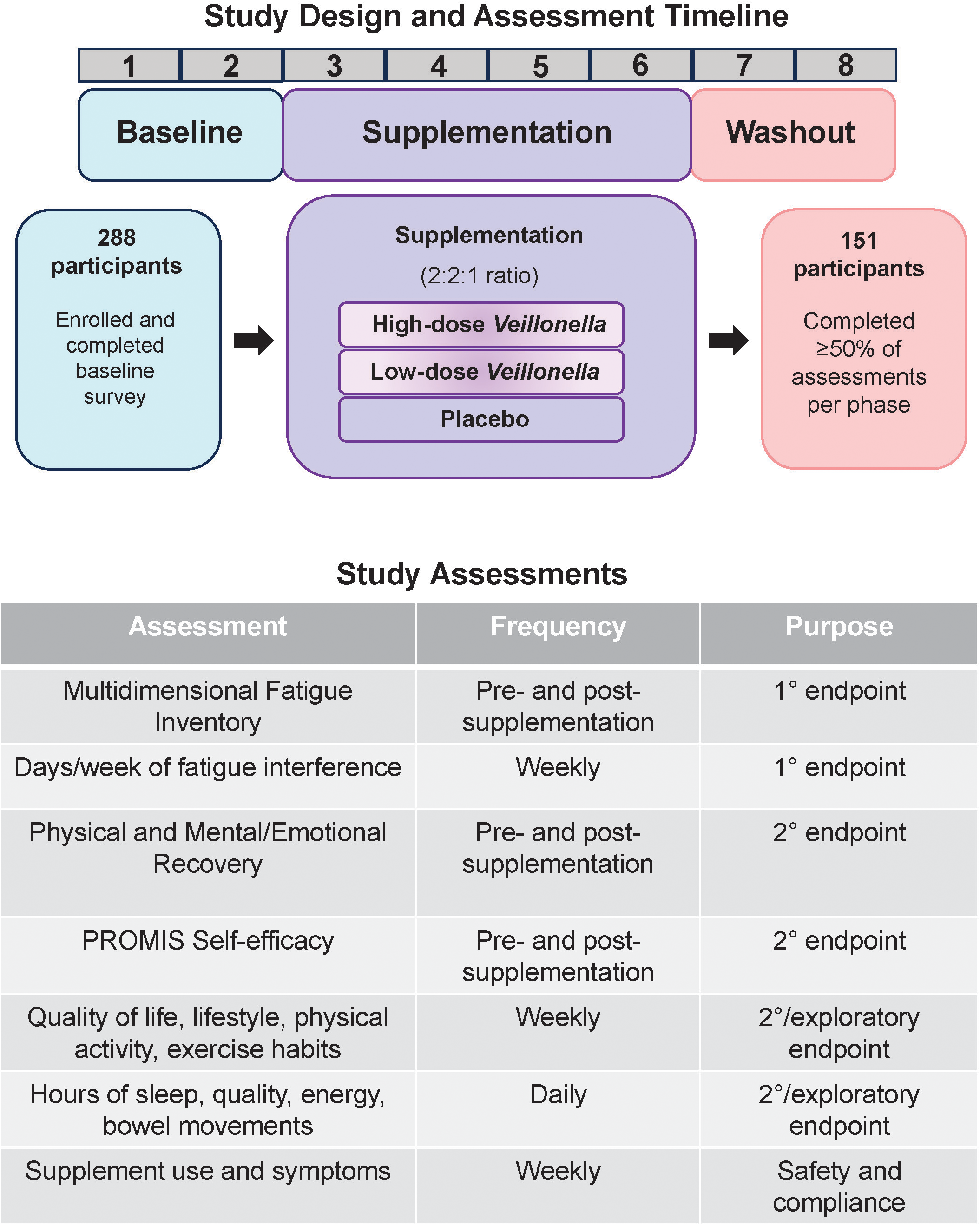
Study design, timeline, and assessment schedule for the randomized controlled trial of *Veillonella atypica* supplementation. Participants were randomized 2:2:1 to high-dose V. atypica (1×10¹⁰ CFU), low-dose V. atypica (5×10⁹ CFU), or placebo. The study comprised three phases: baseline (weeks 1-2), supplementation (weeks 3-6), and washout (weeks 7-8).

At enrollment, 288 participants provided baseline lifestyle and health surveys and were randomized in a 4:4:2 ratio to receive high-dose *V. atypica* (1×10¹⁰ CFU), low-dose *V. atypica* (5×10⁹ CFU), or placebo. Capsules were mailed directly to participants for self-administration once daily. Study activities—including daily assessments of sleep, energy, and bowel movements, and weekly surveys of fatigue interference, physical activity, mood, and general health—were completed remotely through the CHLOE app. Participants also completed validated questionnaires at baseline and post-supplementation: the Multidimensional Fatigue Inventory (MFI-20) and PROMIS General Self-Efficacy survey.

Of the 288 enrolled participants, 151 met compliance criteria (≥50% of surveys completed in each of the three study phases), forming the analytic cohort (**Figure 1**). Preliminary analyses revealed highly similar responses between the two *V. atypica* dose groups—consistent with the modest two-fold dose difference—and these arms were therefore pooled for subsequent analyses (referred to as “*V. atypica*” henceforth). The mean age was 46.7 ± 12.1 years, and 38.4% were female. Baseline demographics, including age and sex distribution, were balanced across treatment arms (**Table 1**).

**Table 1.** Baseline demographic and clinical characteristics of study participants. Demographic and baseline characteristics for participants randomized to *Veillonella atypica* (pooled high-dose and low-dose) or placebo who completed ≥50% of assessments per study phase (n=151). Continuous variables presented as mean ± standard deviation; categorical variables as n (%). Age, sex, body mass index (BMI), baseline physical activity level, and baseline fatigue scores were balanced between treatment arms. No statistically significant differences were observed between groups (age: *p*=0.48, t-test; sex: *p*=0.71, χ² test; other comparisons as indicated in table).

| Participant demographics and baseline characteristics |  |  |  |  |  |
| --- | --- | --- | --- | --- | --- |
| Values are N or n (%); age shown as mean ± SD. |  |  |  |  |  |
|  |  | High | Low | Placebo | Overall |
| Sample size | N | 57 | 66 | 28 | 151 |
| Demographics | Age, years (mean ± SD) | 47.0 ± 11.8 | 44.9 ± 12.0 | 50.2 ± 12.9 | 46.7 ± 12.1 |
| Sex | Female | 22 (38.6%) | 27 (40.9%) | 9 (32.1%) | 58 (38.4%) |
|  | Male | 35 (61.4%) | 39 (59.1%) | 19 (67.9%) | 93 (61.6%) |
|  | Other / NA | 0 (0.0%) | 0 (0.0%) | 0 (0.0%) | 0 (0.0%) |
| Lifestyle | Dedicated To Fitness, Moderate Exercise 3-5 Days Per Week | 27 (47.4%) | 27 (40.9%) | 15 (53.6%) | 69 (45.7%) |
|  | Exercise Addict, Moderate To High Intensity Exercise Almost Every Day | 20 (35.1%) | 21 (31.8%) | 4 (14.3%) | 45 (29.8%) |
|  | Occasional Exerciser, Light Exercise 1-2 Days Per Week | 3 (5.3%) | 11 (16.7%) | 7 (25.0%) | 21 (13.9%) |
|  | Weekend Warrior, Moderate To High Intensity Exercise 1-2 Days Per Week, Light To Moderate Exercise On Other Days | 4 (7.0%) | 5 (7.6%) | — | 9 (6.0%) |
|  | Sedentary, Exercise Less Than Once Per Week | 3 (5.3%) | 2 (3.0%) | 2 (7.1%) | 7 (4.6%) |
| Athlete status | Athlete in training (Yes) | 2 (3.5%) | 4 (6.1%) | 0 (0.0%) | 6 (4.0%) |
| Race / Ethnicity | White / Caucasian | 44 (77.2%) | 58 (87.9%) | 26 (92.9%) | 128 (84.8%) |
|  | Hispanic / Latino | 6 (10.5%) | 5 (7.6%) | 0 (0.0%) | 11 (7.3%) |
|  | Asian / Pacific Islander | 8 (14.0%) | 4 (6.1%) | 4 (14.3%) | 16 (10.6%) |
|  | Black / African American | 1 (1.8%) | 1 (1.5%) | 1 (3.6%) | 3 (2.0%) |
|  | Multiracial / Biracial | 2 (3.5%) | 3 (4.5%) | 0 (0.0%) | 5 (3.3%) |
|  | Native American / Alaskan Native | 1 (1.8%) | 3 (4.5%) | 3 (10.7%) | 7 (4.6%) |
|  | Other (Not Listed) | 2 (3.5%) | 0 (0.0%) | 0 (0.0%) | 2 (1.3%) |

Race/ethnicity categories are not mutually exclusive; percentages may sum to >100%.

#### Safety and Tolerability

Participants reported adverse events (AEs) via the CHLOE app and direct investigator contact. Overall, AE reporting rates were lower for *V. atypica* compared to placebo: 71.4% of placebo participants reported at least one AE versus 61.8% of *V. atypica* participants (relative risk 0.75; 95% CI 0.60 to 0.94; χ² p=0.019; **Supplementary Figure 1A**). However, this difference may have been driven by a baseline imbalance (21.4% vs 10.6% reporting AEs during weeks 1-2 prior to supplementation) and convergent reporting patterns during active treatment (67.9% vs 57.7% during weeks 3-6). When assessed by weekly survey observations, AE reporting rates were 31.4% for placebo and 23.5% for *V. atypica* (**Supplementary Figure 1A**). The temporal pattern of AE reports was similar across treatment arms, with reporting increasing during early supplementation (weeks 3–5) and declining during washout (**Supplementary Figure 1B**).

The most frequently reported AEs in both groups were gastrointestinal, including gas, increased bowel movements, unusual stool texture, diarrhea, constipation, and bloating (**Supplementary Figure 1C**). When specific AE categories were examined individually, no statistically significant differences were observed between treatment arms (Fisher’s exact tests: all *p* > 0.20). The proportion of participants who reported stopping supplementation was similar between groups (placebo: 8.8%; *V. atypica*: 10.3%). Unsolicited non-serious events included isolated reports of kidney pain (n=2, self-limited), heart palpitations (n=1), rash (n=1), weight gain (n=1), and dry eye (n=1). No serious adverse events were reported. The observed safety profile is consistent with probiotic supplementation, characterized by predominantly mild, transient gastrointestinal symptoms that did not result in differential discontinuation between treatment arms.

#### Weekly Longitudinal Assessments Reveal Divergent Trajectories

To capture temporal dynamics beyond pre/post comparisons, participants completed weekly surveys throughout the 8-week study assessing fatigue interference, physical activity, mood, recovery, digestive health, and overall quality of life. Among these endpoints, two showed significant treatment-specific effects: **fatigue interference** (“How many days in the prior week did fatigue interfere with everyday activities?”) and **physical activity hours** (“How many total hours did you engage in physical activity this week?”). We employed Analysis of Covariance (ANCOVA) models adjusting for individual baseline levels (calculated as the mean of the first two study weeks) to account for pre-existing differences and increase statistical power.

#### Fatigue Interference

Visual inspection of weekly trajectories revealed an early, transient reduction in fatigue interference in both treatment arms during the first two weeks of supplementation (**Figure 2A**), consistent with initial placebo or study-engagement effects. However, the two groups diverged markedly thereafter: while placebo participants’ scores gradually returned toward baseline levels across weeks 3–8, *V. atypica* participants sustained lower fatigue interference throughout both the supplementation and washout phases.

**Figure 2.**
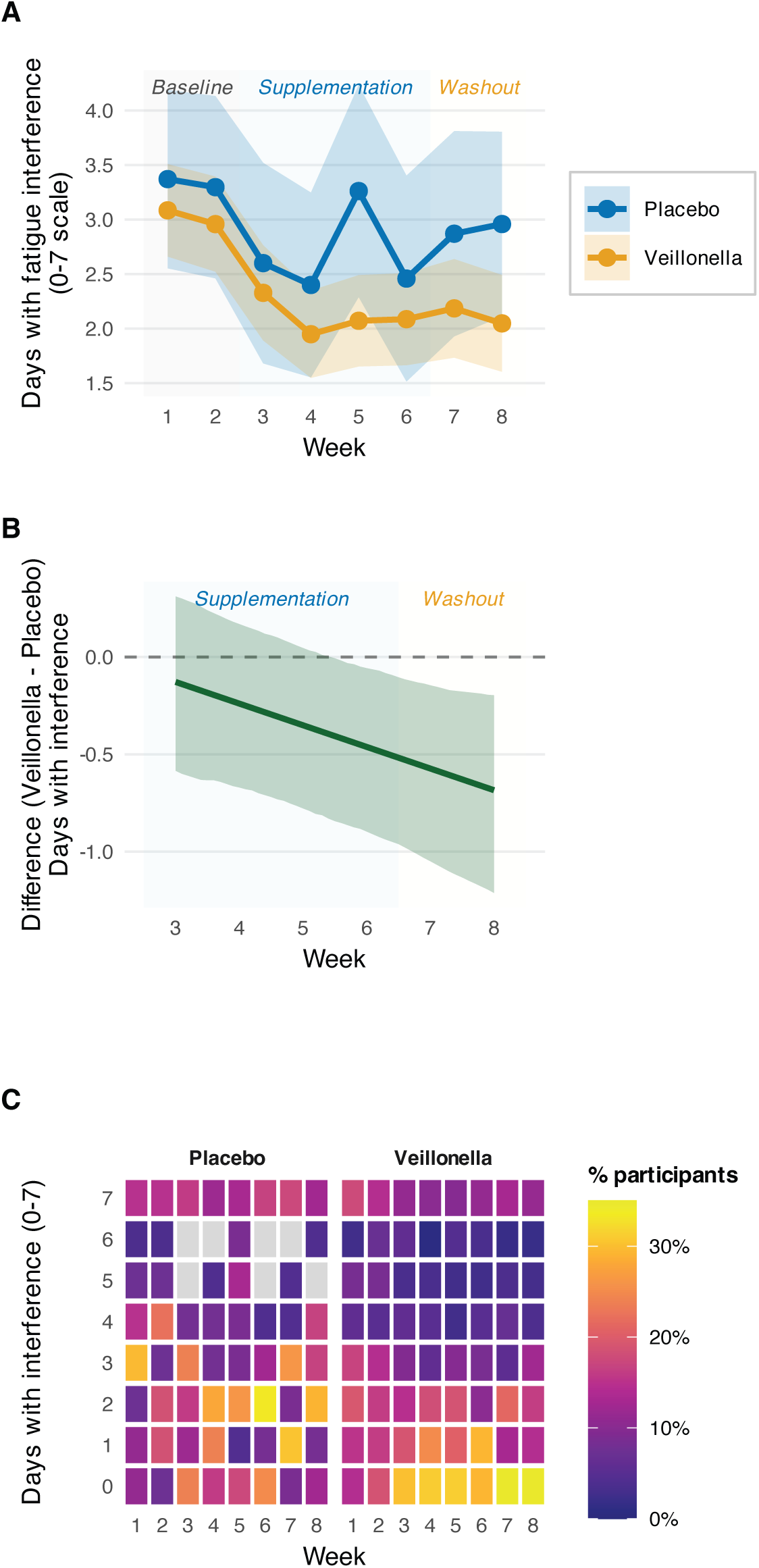
*Veillonella atypica* supplementation reduces fatigue interference through sustained trajectory improvement. (**A**) Weekly trajectories of fatigue interference (0–7 days/week scale) for placebo (blue) and *V. atypica* (orange) groups across the 8-week study. Points represent group means; ribbons show 95% confidence intervals calculated from standard errors. Both groups show initial decline during early supplementation (weeks 2–3), consistent with study engagement effects, but diverge thereafter with placebo returning toward baseline while *V. atypica* maintains reduced interference. (**B**) Model-predicted treatment effect (*V. atypica* − placebo) from linear mixed-effects ANCOVA adjusting for individual baseline values (mean of weeks 1–2). Predictions generated at the grand mean baseline using a fine time grid (0.1-week increments) for smooth trajectories; bootstrap 95% confidence intervals (1,000 iterations) shown as shaded ribbon. Negative values indicate lower fatigue interference in the *V. atypica* group. The progressive divergence reaches approximately −0.6 days by week 6. Treatment×week interaction: β = −0.097 days/week (95% CI −0.175 to −0.018), *p* = 0.016. (**C**) Distribution heatmaps showing the percentage of participants reporting each fatigue interference level (0–7 days) at each week, separately by treatment arm. Tiles are colored using the viridis scale (option C); darker colors indicate higher percentages. Missing cells (NA values representing levels with zero observations) displayed in gray. During supplementation and washout (weeks 2–7), the *V. atypica* group shows marked concentration at lower interference levels (0–2 days), visible as dark tiles in the bottom rows, whereas placebo shows more dispersed distribution centered at 3–5 days. This distributional shift indicates that the mean reduction reflects a genuine population-level change rather than extreme responders. Background shading indicates study phases: gray = Baseline (weeks 1–2), blue = Supplementation (weeks 3–6), yellow = Washout (weeks 7–8). *V. atypica*, n = 119; Placebo, n = 27. ANCOVA model formula: interference ∼ baseline_interference + treatment × week_post_baseline + (1|participant_id), fitted to weeks 3–8 data.

Linear mixed-effects ANCOVA, adjusting for each participant’s baseline fatigue interference (mean of weeks 1–2) and including random intercepts for participants, confirmed a significant **treatment×week interaction** (β = −0.097 days/week; 95% CI −0.175 to −0.018; *p* = 0.016). This interaction indicates that the *V. atypica* group exhibited a significantly faster rate of decline in fatigue interference relative to placebo over the post-baseline period (weeks 3–8). Model-predicted treatment effects (**Figure 2B**) illustrate the progressive divergence between groups, with the difference reaching approximately −0.6 days by week 6, corresponding to roughly one fewer day per week where fatigue interfered with daily activities.

Analysis of the distributional shift (**Figure 2C**) revealed that this mean reduction reflected a genuine shift in the response distribution rather than isolated changes in a few individuals. During the supplementation phase, the *V. atypica* group showed increasing concentration at lower interference levels (0–2 days/week), whereas the placebo group maintained a more dispersed distribution centered at higher values (3–5 days/week). This pattern persisted into the washout phase (weeks 7–8), suggesting durability beyond active supplementation.

At baseline (weeks 1–2), the groups reported similar mean fatigue interference (placebo: 3.40 ± 0.32 days/week; *V. atypica*: 3.08 ± 0.11 days/week; *p* = 0.34), confirming successful randomization. Baseline fatigue was a strong predictor of post-baseline interference (β = 0.88; 95% CI 0.84 to 0.92; *p* < 2×10⁻¹⁶), underscoring the importance of covariate adjustment in this analysis.

#### Physical Activity Hours

A complementary pattern emerged for self-reported weekly physical activity hours (**Figure 3A**). While both groups reported comparable baseline activity levels (placebo: 7.92 ± 0.94 h/week; *V. atypica*: 9.23 ± 0.57 h/week; *p* = 0.22), participants randomized to *V. atypica* exhibited a progressive increase in weekly activity hours across the supplementation phase that was not observed in the placebo group. Physical activity values were capped at 40 hours/week to address extreme outliers (values >40 h/week were considered implausible for this population).

**Figure 3.**
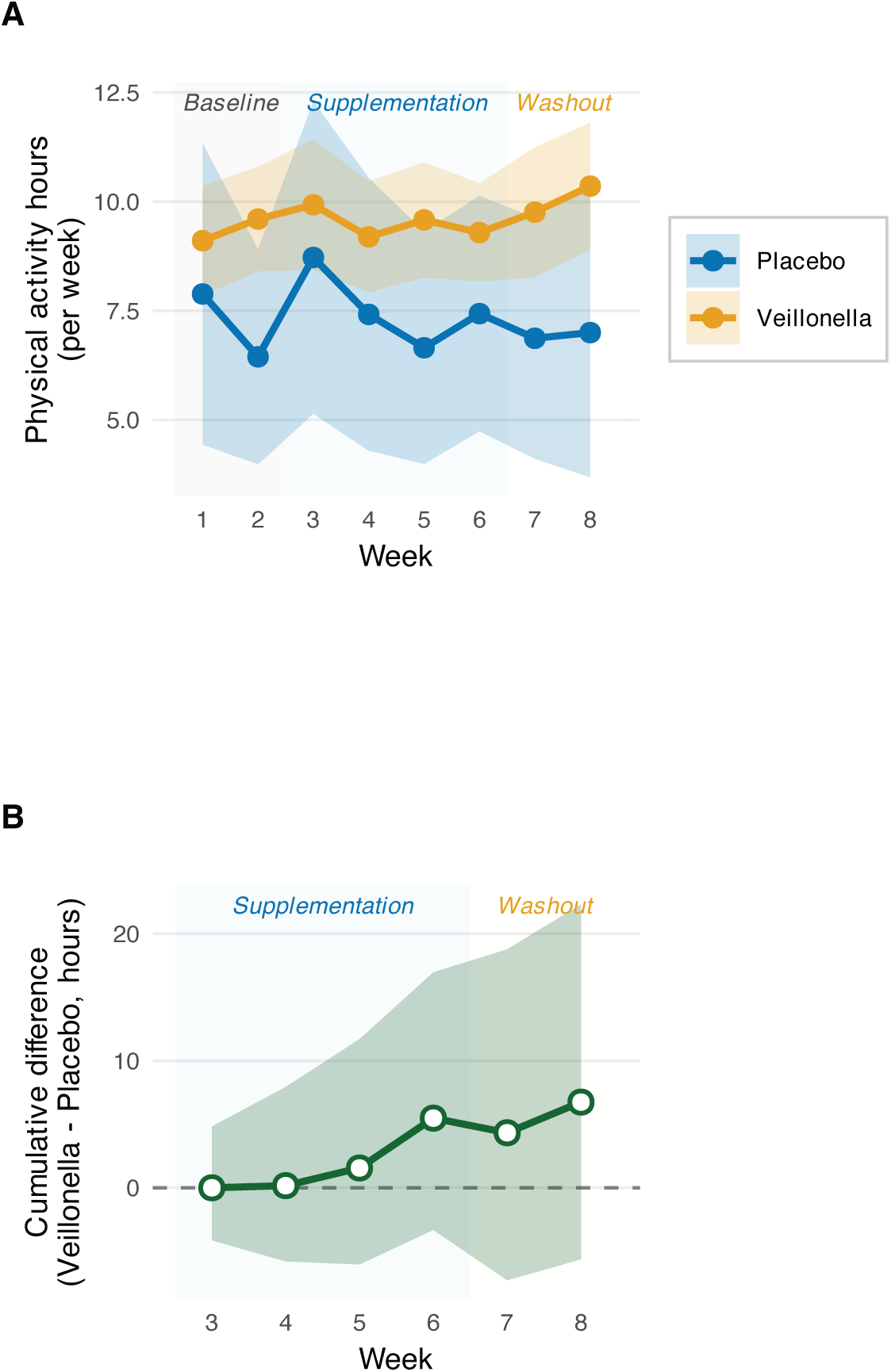
*Veillonella atypica* supplementation increases voluntary physical activity engagement. (**A**) Weekly trajectories of self-reported physical activity hours for placebo (blue) and *V. atypica* (orange) groups across the 8-week study. Points represent group means; ribbons show 95% confidence intervals calculated from standard errors. Baseline activity levels (weeks 1–2) are comparable between groups, but *V. atypica* participants exhibit progressive increases during supplementation (weeks 3–6) that persist into washout (weeks 7–8), whereas placebo shows minimal change. Physical activity values capped at 40 h/week to address implausible extreme outliers. (**B**) Model-predicted cumulative treatment effect (*V. atypica* − placebo) normalized to zero at week 2 (supplementation onset). This metric captures the accumulating benefit over time by summing weekly changes from baseline for each participant, then computing group-level cumulative differences. Bootstrap 95% confidence intervals (2,000 iterations) shown as shaded ribbon. The upward trajectory during supplementation demonstrates sustained engagement enhancement, with the cumulative difference reaching approximately 10– 12 hours by week 7 (equivalent to ∼2 additional hours per week averaged across the 6-week post-baseline period). Treatment×week interaction from ANCOVA adjusting for individual baseline values: β = +0.357 h/week (95% CI +0.031 to +0.684), *p* = 0.032. Background shading indicates study phases: gray = Baseline (weeks 1– 2), blue = Supplementation (weeks 3–6), yellow = Washout (weeks 7–8). *V. atypica*, n = 119; Placebo, n = 27. ANCOVA model formula: activity_hours ∼ baseline_activity + treatment × week_post_baseline + (1|participant_id), fitted to weeks 3–8 data with 40 h/week cap applied.

Mixed-effects ANCOVA adjusting for baseline physical activity hours confirmed a significant **treatment×week interaction** (β = +0.357 h/week; 95% CI +0.031 to +0.684; *p* = 0.032). Placebo participants showed minimal change in activity over time (estimated slope ≈ −0.08 h/week, not significant), whereas *V. atypica* participants exhibited a progressive upward trajectory (estimated slope ≈ +0.28 h/week, *p* = 0.012). Model-predicted treatment effects (**Figure 3B**) demonstrate that by week 6, the cumulative difference approached approximately 2 additional hours per week of physical activity in the *V. atypica* group relative to placebo.

Notably, this increase in activity hours occurred without accompanying increases in reported adverse events, injuries, or gastrointestinal complaints (see Safety and Tolerability section), supporting the tolerability of higher activity engagement. The benefit persisted into the washout period (weeks 7–8), with *V. atypica* participants maintaining elevated activity levels relative to their baseline, consistent with the sustained reduction in fatigue interference observed during the same period.

Other weekly survey measures—including mood, recovery, digestive health, general health, quality of life, physical activity frequency and intensity ratings, and energy levels—showed no significant treatment×week interactions (all *p* > 0.15; **Supplementary Figure 2**), indicating specificity of effects to fatigue interference and physical activity hours rather than global improvement across all domains. This pattern supports a targeted mechanism related to exercise motivation and perceived effort rather than diffuse, non-specific wellness effects.

#### Multidimensional Fatigue Inventory

To assess fatigue using a validated, multidimensional instrument, participants completed the Multidimensional Fatigue Inventory (MFI-20) at baseline (week 0) and at the end of the supplementation phase (week 6). The MFI-20 comprises five subscales—General Fatigue, Physical Fatigue, Reduced Activity, Reduced Motivation, and Mental Fatigue—each scored from 4 (low fatigue) to 20 (high fatigue). These subscales sum to a Total MFI score ranging from 20 (minimal fatigue) to 100 (maximal fatigue).

Both treatment groups—*V. atypica* and placebo—exhibited substantial reductions in Total MFI scores from baseline to post-supplementation (**Supplementary Figure 3**). However, linear mixed-effects models demonstrated that these improvements were largely comparable between groups, with no evidence of differential treatment effects. Placebo participants improved from a baseline Total MFI of 51.0 ± 2.9 to 44.8 ± 3.0 post-supplementation (mean change −6.2 ± 1.6), while *V. atypica* participants improved from 50.8 ± 1.7 to 43.8 ± 1.7 (mean change −7.0 ± 1.2). A linear mixed-effects model with treatment, time, and their interaction as fixed effects, and random intercepts for participants, revealed a significant within-subject time effect (F=27.4, *p*=9.8×10⁻⁷) but no significant treatment×time interaction (F=0.19, *p*=0.66), indicating parallel improvement trajectories. The between-group difference in change scores was −0.8 points (95% CI −4.2 to +2.6; *p*=0.66).

Analysis of all individual MFI subscales revealed a consistent pattern, with robust time effects but no between-group differences in trajectories. Complete statistics for all subscales are provided in **Supplementary Table 1**.

#### Daily Assessments of Sleep Quality and Energy

Participants completed brief daily surveys throughout the study capturing sleep duration (hours), sleep quality (5-point Likert scale: 1=very poor, 5=very good), self-reported energy level (binary: high energy day yes/no), and bowel movements (count). These high-frequency measures provided insight into day-to-day variability and potential mechanisms linking *V. atypica* to behavioral changes.

#### Sleep Quality

Daily sleep quality assessments revealed treatment-specific patterns that emerged during the washout period (**Figure 4A**). Linear mixed models examining sleep quality across the three study phases (baseline, supplementation, washout) showed no treatment differences during baseline (mean difference +0.17 units; 95% CI −0.11 to +0.44; *p*=0.22) or supplementation (*p*=0.15), but a significant difference emerged during washout: *V. atypica* participants reported higher sleep quality than placebo (mean difference +0.31 units; 95% CI +0.04 to +0.58; *p*=0.024).

**Figure 4.**
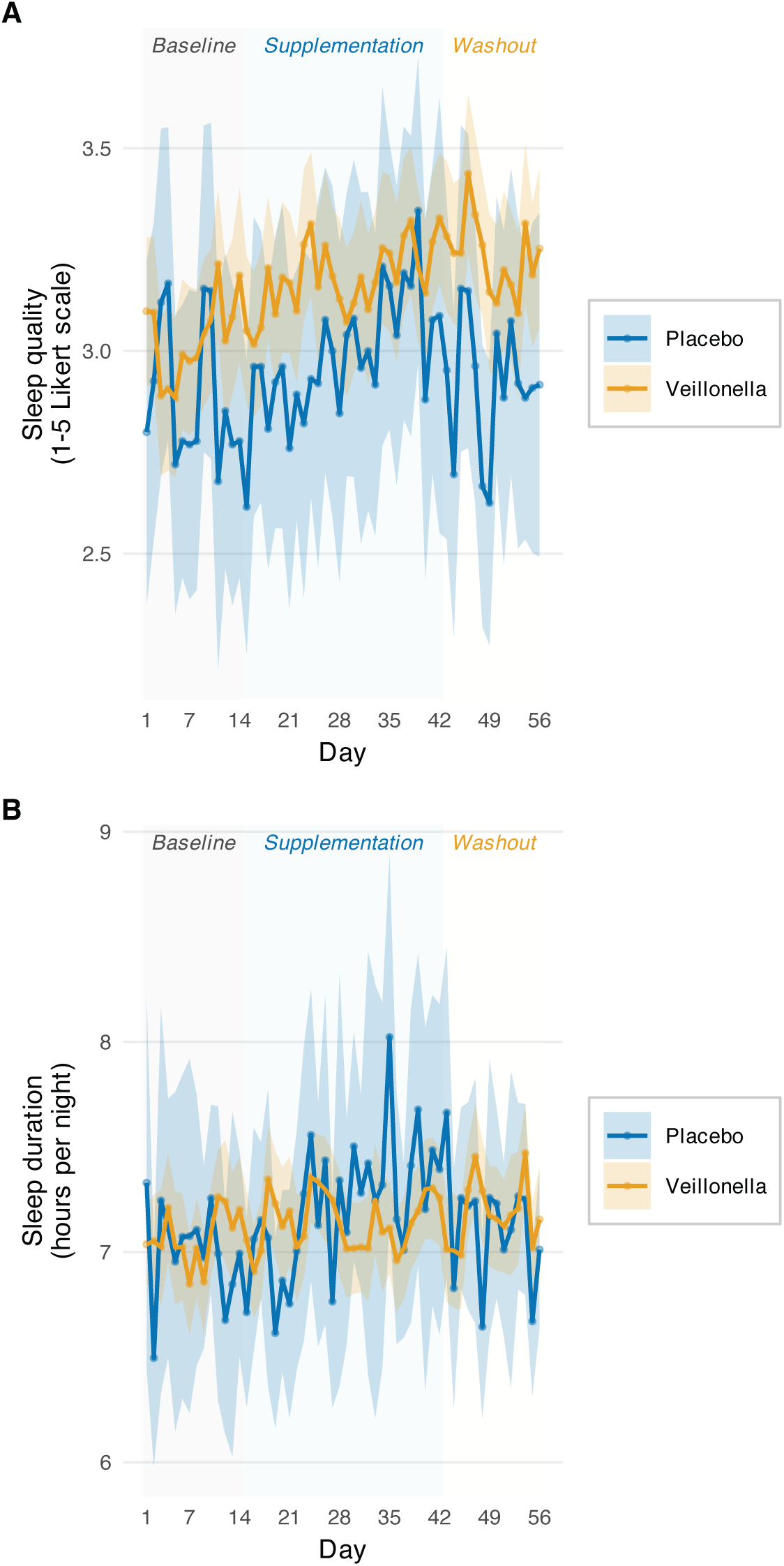
Daily sleep quality trajectories show *Veillonella atypica*-specific improvement. (**A**) Sleep quality ratings (1–5 Likert scale: 1=very poor, 5=very good) across the full study period (days 0–56) for placebo (blue) and *V. atypica* (orange) groups. Individual participant data shown as semi-transparent points; smoothed group trajectories (LOESS) shown as bold lines with 95% confidence bands. The *V. atypica* group shows progressively higher sleep quality during supplementation and washout phases, with the largest divergence during washout (days 42–56). Mixed-effects models revealed no treatment differences during baseline (*p*=0.22) or supplementation (*p*=0.15), but significant differences emerged during washout (mean difference +0.31 units, 95% CI +0.04 to +0.58, *p*=0.024). Difference-in-differences analysis comparing baseline to washout confirmed differential change between groups (*p*=0.032). (**B**) Sleep duration (hours per night) across the study period. No treatment×period interactions were detected for either supplementation (*p*=0.61) or washout (*p*=0.34) periods. Mean sleep duration: placebo 7.1±0.1 h/night; *V. atypica* 7.2±0.1 h/night. The dissociation between improved sleep quality (panel A) without increased duration suggests effects on sleep architecture or subjective restorativeness rather than sleep extension. Vertical shaded regions demarcate study phases: Baseline (gray, days 0–13), Supplementation (blue, days 14–41), and Washout (yellow, days 42–56). *V. atypica*, n=119 contributing 5,845 daily observations; Placebo, n=27 contributing 1,247 daily observations. Mixed model formula: outcome ∼ treatment × period + (1|participant_id), fitted separately for baseline vs. supplementation and baseline vs. washout comparisons.

To test whether this washout difference reflected differential change from baseline, we conducted a difference-in-differences (DID) analysis comparing baseline (days 0–13) to washout (days 42–56). The **treatment×period interaction** was significant (*p*=0.032), confirming that the groups diverged specifically between these phases. Placebo participants’ mean sleep quality declined from baseline (3.06 ± 0.05) to washout (2.89 ± 0.06; change: −0.17 ± 0.08), whereas *V. atypica* participants maintained stable quality (baseline: 3.07 ± 0.03; washout: 3.09 ± 0.04; change: +0.02 ± 0.05). The between-group difference in these changes corresponds to approximately one-fifth of the scale range.

Trajectory modeling with treatment×period×time interactions revealed that sleep quality dynamics differed between groups across study phases (*p*=0.024 for supplementation interaction). A parallel DID analysis comparing baseline to the supplementation period (days 14–41) showed no significant treatment×period interaction (*p*>0.15), indicating that group divergence occurred specifically during washout rather than during active treatment. This temporal pattern—preservation of sleep quality during washout rather than improvement during supplementation—suggests effects that are delayed until several weeks after begining active bacterial supplementation and persist beyond the period of supplementation.

Importantly, sleep quality improvements were **not accompanied by changes in sleep duration** (**Figure 4B**). Mixed models revealed no treatment×period interactions for sleep duration in either the supplementation (*p*=0.61) or washout (*p*=0.34) periods. Both groups reported similar sleep duration throughout (placebo: 7.1 ± 0.1 h/night; *V. atypica*: 7.2 ± 0.1 h/night). This dissociation—maintained sleep quality without increased duration—suggests effects on sleep architecture, sleep fragmentation, or subjective restorativeness rather than sleep extension.

#### Daily Energy Levels

Participants reported whether each day was a “high energy” day (binary outcome). The proportion of high-energy days was consistently higher in the *V. atypica* group across the study, with trends becoming more pronounced during supplementation and washout (**Supplementary Figure 4A**).

Using mixed-effects logistic regression with treatment, period (baseline/supplementation/washout), and their interaction as predictors, the odds of reporting a high-energy day trended higher in the *V. atypica* group during washout (OR=2.36; 95% CI 0.93 to 6.02; *p*=0.071), though this did not reach conventional statistical significance. During supplementation, the odds ratio was more modest (OR=1.48; 95% CI 0.74 to 2.94; *p*=0.27). Baseline proportions were similar (placebo: 62% high-energy days; *V. atypica*: 65%; *p*=0.58).

The trend toward increased high-energy days during washout parallels the sleep quality findings and may reflect shared underlying mechanisms or causal relationships (e.g., better sleep → more energy). However, the marginal statistical significance (*p*=0.071) and lack of effect during active supplementation warrant cautious interpretation.

#### Bowel Movements

Mean daily bowel movements did not differ between treatment arms at any phase of the study (**Supplementary Figure 4B**). Mixed models revealed no treatment×period interactions for either supplementation (*p*=0.88) or washout (*p*=0.72) periods. Both groups reported similar bowel movement frequency throughout (placebo: 1.93 ± 0.08 per day; *V. atypica*: 1.98 ± 0.05 per day). This absence of gastrointestinal effects is consistent with the adverse event profile and supports the tolerability of *V. atypica* supplementation.

### VOLUNTARY WHEEL RUNNING IN MICE

#### Study Design and Colonization Verification

To establish biological plausibility, test cross-species generalizability, and investigate potential neural mechanisms, we profiled voluntary wheel running in C57BL/6J male mice randomized to receive PBS (n=8), *V. atypica* (n=8), or *Lactobacillus johnsonii* (n=8) delivered in peanut butter (**Figure 5A**). *L. johnsonii* served as an active control: this bacterium produces lactate through fermentation rather than consuming it, thereby testing specificity for lactate-utilizing metabolism. Mice were single-housed with continuous access to running wheels, and distance was logged automatically throughout the study.

**Figure 5.**
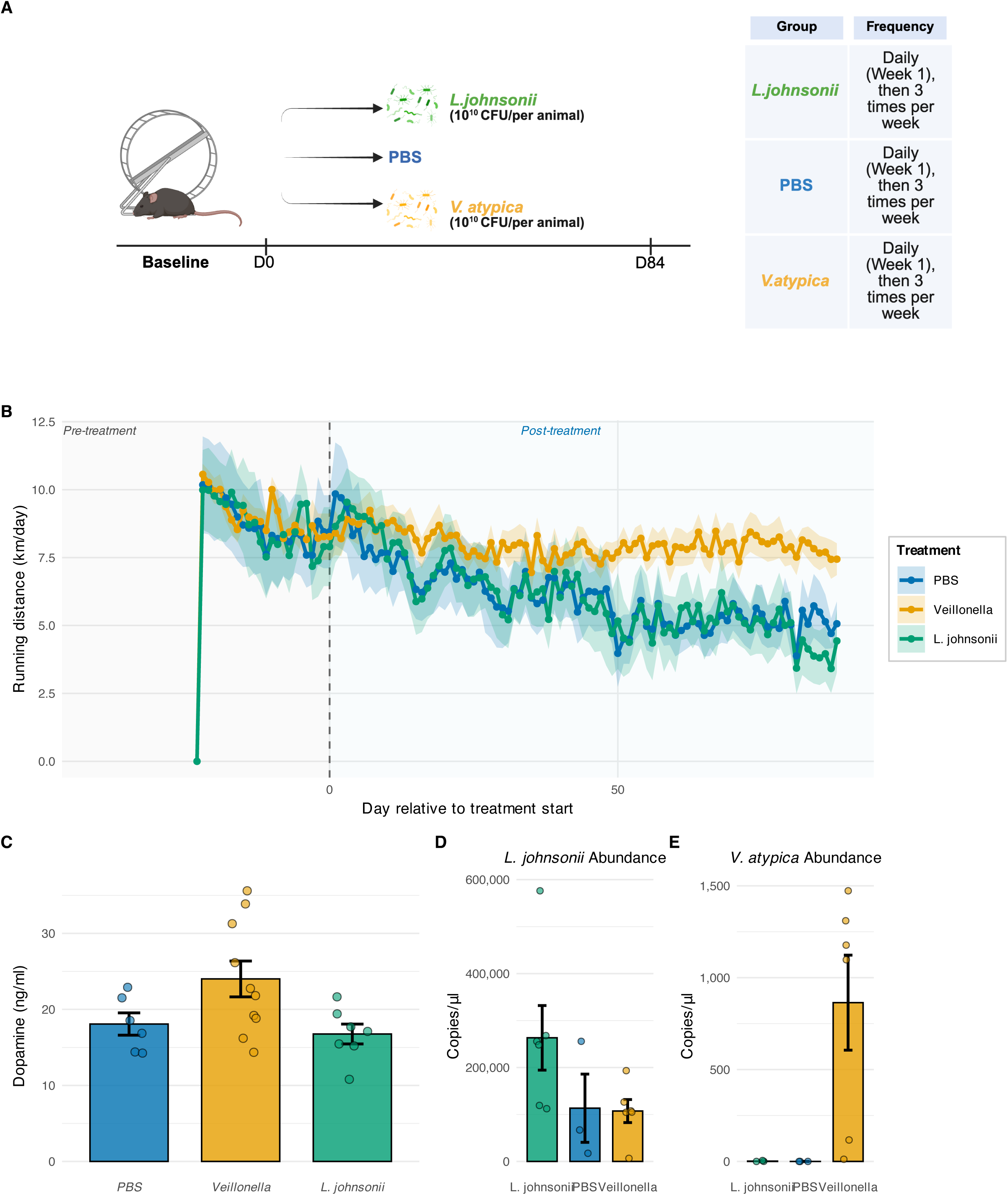
Voluntary wheel running in mice is maintained by *Veillonella atypica* supplementation with elevated striatal dopamine. (**A**) Experimental schematic. C57BL/6J male mice (4 months old, n=8 per group) were randomized to receive PBS (vehicle control), *Veillonella atypica*, or *Lactobacillus johnsonii* (lactate-producing control) delivered in peanut butter. Following ∼3-week acclimation with ad libitum wheel access, supplementation began (Day 0) and continued for 50 days, followed by 34-day washout. Mice were single-housed with continuous automated logging of wheel revolutions. Fecal samples were collected weekly for colonization monitoring. **(B)** Voluntary running wheel activity over time, showing *Veillonella*-supplemented mice maintain higher running distances post-treatment compared to PBS and *L. johnsonii* controls (n = mice per group, mean ± SEM). Shaded regions indicate pre-treatment (gray) and post-treatment (light blue) periods. **(C)** Striatal dopamine concentrations show elevation in *Veillonella* group compared to controls, suggesting a neurochemical mechanism for enhanced exercise motivation (bars show mean ± SEM, individual data points overlaid). **(D-E)** Bacterial colonization at Day 50 confirms successful engraftment of both *L. johnsonii* and *V. atypica* in their respective treatment groups with minimal cross-contamination (mean ± SEM with individual data points). These findings support a gut-brain axis mechanism whereby *V. atypica* colonization influences dopaminergic signaling to enhance voluntary physical activity.

Following a three-week acclimation period to establish stable baseline running, supplementation began (Day 0) and continued for 50 days, followed by a 34-day washout period (84 days total post-baseline monitoring). Quantitative PCR analysis of fecal DNA confirmed successful colonization: by Day 50, *V. atypica* was detected at high levels exclusively in the *V. atypica* group (8.4×10² copies/μL versus <10 copies/μL in controls, *p*<0.001), while *L. johnsonii* was elevated only in its designated group (2.5×10⁵ versus <10⁴ copies/μL in controls, *p*=0.041; **Figure 5D & 5E**). These results confirm arm-specific colonization without cross-contamination.

#### *V. atypica* Maintains Voluntary Running Over Time

All groups exhibited comparable baseline running levels prior to supplementation (PBS: 9.1 ± 0.8 km/day; *V. atypica*: 9.6 ± 0.9 km/day; *L. johnsonii*: 9.8 ± 0.7 km/day; *p*=0.81). However, trajectories diverged markedly following supplementation initiation (**Figure 5B**). PBS and *L. johnsonii* groups showed progressive declines in running distance—a well-documented phenomenon in aging mice and extended voluntary running paradigms— whereas the *V. atypica* group maintained stable running throughout the 84-day monitoring period.

Linear mixed-effects modeling with group, day, and group×day interaction as fixed effects, plus random intercepts and slopes for individual mice, revealed a highly significant interaction (*F*=13.31, *p*=2.1×10⁻⁴), indicating different rates of change across groups. The *V. atypica* trajectory was essentially flat (slope −0.0077 km/day/day, *p*=0.19), while both control groups declined significantly (PBS: −0.0394 km/day/day, *p*<0.001; *L. johnsonii*: −0.0497 km/day/day, *p*<0.001). Pairwise comparisons confirmed that *V. atypica* slopes differed from both PBS (*p*=0.003) and *L. johnsonii* (*p*<0.001), while the two control groups did not differ from each other (*p*=0.56). Quantitatively, during the final five weeks of monitoring (Days 49–84), *V. atypica*-treated mice ran approximately 2.86 km/day more than PBS controls (*p*=2.2×10⁻⁴). This sustained behavioral difference represents a preservation of voluntary exercise engagement rather than enhancement beyond baseline levels.

#### Striatal Dopamine: Preliminary Mechanistic Insight

To investigate potential neural mechanisms linking *V. atypica* to sustained voluntary activity, we quantified dopamine concentrations in the dorsal striatum at study endpoint (Day 84) using ELISA (**Figure 5C**). The dorsal striatum—encompassing caudate and putamen—is a key node in motor and motivational circuits, and striatal dopamine signaling has been implicated in both voluntary exercise initiation and sustained engagement in prior studies.

Dorsal striatal dopamine concentrations differed across groups by one-way ANOVA (*F*=3.46, *p*=0.05). Mice receiving *V. atypica* exhibited approximately 30% higher dopamine levels (23.4 ± 2.1 ng/mg tissue) compared to PBS controls (17.8 ± 1.2 ng/mg, *p*=0.03) and a similar magnitude elevation relative to *L. johnsonii* (16.2 ± 1.7 ng/mg, *p*=0.08). The two control groups did not differ from each other (*p*=0.68). While this finding provides preliminary evidence consistent with dopaminergic modulation, several important caveats temper interpretation: the overall ANOVA reached exactly *p*=0.05, measurements were conducted at a single endpoint rather than dynamically during running, and whole-tissue homogenates lack the regional resolution to distinguish specific striatal subregions with distinct functional roles. These limitations are addressed further in the Discussion.

## DISCUSSION

In a randomized controlled trial of 151 healthy adults, *Veillonella atypica* supplementation reduced fatigue interference and increased voluntary physical activity over 8 weeks, as assessed by weekly longitudinal surveys. Parallel mouse studies showed that *V. atypica* maintained voluntary wheel running across 12 weeks, whereas PBS and *Lactobacillus johnsonii* controls exhibited significant declines. Mice receiving *V. atypica* showed elevated striatal dopamine concentrations at study endpoint, consistent with recent evidence that the gut microbiome regulates exercise motivation through dopaminergic signaling [18]. Together, these findings suggest that *V. atypica* influences voluntary physical activity through gut-brain mechanisms, though the precise pathways require further validation.

We previously demonstrated that *V. atypica*, isolated from elite marathon runners, enhances treadmill running time to exhaustion in mice through lactate-to-propionate metabolism [12]. However, that work employed forced exercise paradigms that measure physiological capacity under compulsion rather than voluntary engagement. The distinction is clinically important: most sedentary individuals are physically capable of moderate activity but lack motivation to initiate and sustain exercise [5]. Interventions targeting exercise motivation must therefore address voluntary behavior governed by central neural circuits, including striatal dopamine signaling [15], rather than peripheral performance capacity alone. Our findings that *V. atypica* increases self-reported activity hours in humans and maintains voluntary wheel running in mice extend previous forced-exercise results to motivationally relevant outcomes.

The observed behavioral effects are consistent with a pathway in which *V. atypica* converts exercise-derived lactate to propionate, which crosses the blood-brain barrier and modulates striatal dopamine. During exercise, muscle lactate production increases substantially and lactate is transported into the gut lumen, where it becomes available for microbial metabolism [14]. *Veillonella* species are obligate lactate utilizers that convert lactate to propionate via the succinate pathway [13]. Short-chain fatty acids including propionate cross the blood-brain barrier via monocarboxylate transporters and can affect brain function through multiple mechanisms [20, 21]. Propionate has been shown to increase tyrosine hydroxylase expression in neuronal cell lines [24], protect dopaminergic neurons in Parkinson’s disease models [23], and reduce blood oxygen level-dependent signal in human caudate and nucleus accumbens during reward tasks [22]. Striatal dopamine signaling is necessary for normal levels of voluntary exercise in mice, and dopamine release in ventral striatum increases during running [18]. However, we did not measure propionate concentrations in any compartment—gut, blood, or brain—and thus cannot confirm this proposed mediator. Our striatal dopamine findings, while consistent with this framework, represent single-timepoint measurements in whole-tissue homogenates rather than region-specific or dynamic assessments during behavior. Mechanistic validation through receptor knockout studies, dopamine antagonism, propionate quantification, and real-time neurotransmitter measurements is required to establish causality.

Other mechanisms may contribute to the observed effects. Propionate and other short-chain fatty acids enhance muscle mitochondrial biogenesis through AMPK and PGC-1α signaling [25], which could reduce peripheral fatigue and perceived exertion independent of central motivational effects. Gut microbiota influence brain function through vagal afferent pathways in addition to blood-borne metabolites [26, 27], and propionate may activate vagal neurons expressing FFAR3 receptors. Modulation of tryptophan metabolism and gut serotonin production represents another potential pathway [28]. The reduction in fatigue interference we observed could reflect contributions from multiple systems—central (reduced effort cost perception), peripheral (improved muscle oxidative capacity), or both acting synergistically.

An important methodological finding was that treatment effects emerged in weekly trajectory analyses but not in pre-post assessments of the Multidimensional Fatigue Inventory. Both treatment groups showed substantial MFI improvements from baseline to week 6 (mean reductions 6-7 points), but the trajectories were parallel with no between-group differences (all treatment×time interactions p>0.40). By contrast, weekly assessments of fatigue interference and physical activity hours revealed progressive divergence between groups, with *V. atypica* participants maintaining improvements that gradually returned toward baseline in the placebo group. This pattern suggests that snapshot pre-post designs may miss treatment effects that accumulate gradually and are better captured through repeated measurement. The persistence of behavioral effects into the 2-week washout period raises questions about mechanism durability—whether through continued microbial colonization, stable changes to the broader gut community, behavioral entrainment, or physiological adaptations such as mitochondrial biogenesis that outlast active supplementation.

The improvement in sleep quality (p=0.0039) was a particularly unexpected finding. Sleep duration did not change, suggesting effects on sleep architecture or subjective restorativeness rather than sleep extension. The bidirectional relationships between exercise, sleep, and fatigue warrant further investigation with objective sleep measures and microbiome profiling across all study phases.

Several limitations require acknowledgment. Our prespecified primary endpoint—change in MFI scores from baseline to post-supplementation—showed no treatment differences. Significant effects emerged only in secondary weekly trajectory analyses that were not adjusted for multiple comparisons. While the trajectory findings are biologically plausible, validated in an independent mouse model, and consistent with mechanistic hypotheses, replication in a confirmatory trial with trajectory endpoints as prespecified primary outcomes is necessary to draw more robust conclusions about clinical efficacy. Individual heterogeneity in response was not characterized. Relevant moderators—baseline microbiome composition, host genetics (FFAR3, MCT1, dopamine receptor polymorphisms), exercise history, sex, diet—were not systematically analyzed. Our cohort was 38.4% female and had relatively high baseline activity levels (7-9 hours/week), limiting generalizability to more sedentary populations or those with clinical conditions characterized by fatigue and reduced activity. The study duration would need to be extended significantly beyond 8 weeks to provide information about long-term colonization persistence, behavioral sustainability, and optimal dosing strategies. Finally, mechanistic evidence in support of a gut-brain axis model is preliminary, and would require loss-of-function validation, dynamic measurements during running, and metabolic flux analysis across compartments (feces, plasma, CSF, brain) to become more concretely established.

Future work should focus on mechanistic validation through receptor knockout studies, dopamine antagonists, metabolite quantification including stable isotope tracers, and real-time neurotransmitter measurements during exercise behavior. Responder phenotyping through baseline multi-omics profiling may enable identification of individuals most likely to benefit. Longer-duration studies with intermittent microbiome sampling would address questions of colonization persistence and effect durability. Testing in clinical populations with fatigue-related disorders, metabolic syndrome, or psychiatric conditions affecting motivation would establish therapeutic utility beyond healthy adults. The combination of human behavioral data, cross-species validation in mice, preliminary mechanistic findings consistent with established pathways, and specificity for lactate-utilizing bacteria provides initial evidence that *V. atypica* may represent a viable target for microbiome-based interventions addressing motivational barriers to physical activity, though additional work is required to define mechanisms, optimize implementation, and identify responder populations.

In conclusion, across complementary human and murine experiments, *V. atypica* supplementation was associated with reduced fatigue interference and increased voluntary physical activity, with convergent evidence of sustained behavioral engagement. Preliminary findings of elevated striatal dopamine, integrated with established biochemical pathways and recent demonstrations of microbiome-dopamine-exercise links, support the hypothesis that behavioral effects involve gut-brain signaling to motivational circuits. While mechanistic causality requires definitive validation through loss-of-function experiments and dynamic metabolite measurements, our findings position *V. atypica* as a promising precision microbiome target for addressing motivational barriers to physical activity. In contrast to generic probiotics, *V. atypica* represents mechanism-driven strategy: a microbe selected for specific metabolic capability that becomes active during the behavior of interest, produces a defined neuroactive metabolite with known CNS targets, and modulates a neural circuit established to govern exercise motivation. As the field matures toward mechanistic interventions with defined molecular targets, *V. atypica* and the lactate-propionate-dopamine pathway serve as a model for how gut-brain signaling can be harnessed to influence complex behaviors relevant to the global physical inactivity pandemic.

## MATERIALS & METHODS

### CLINICAL STUDY

#### Study Design and Ethical Approval

This single-center, randomized, single-blind, placebo-controlled trial (ClinicalTrials.gov NCT06141343; Protocol FB004) was reviewed and approved by Advarra Institutional Review Board (Columbia, MD) prior to participant enrollment. The study was conducted in accordance with the Declaration of Helsinki and US Code of Federal Regulations Title 45 Part 46. All participants provided electronic informed consent via the CHLOE application (Consumer Health Learning and Organizing Ecosystem; PeopleScience, Inc.) before any study procedures.

#### Participants

Participants were recruited through email and social media advertisements from August through November 2023. Eligible participants were healthy adults aged 18-65 years, fluent in English, willing to maintain stable diet and exercise habits throughout the study, and able to use a personal smartphone to complete app-based assessments. Key exclusion criteria included: pregnancy or lactation; current or planned antibiotic use; immunosuppressive medications or conditions; chronic illness affecting gut barrier integrity; or conditions contraindicating probiotic supplementation.

#### Randomization and Blinding

Following electronic informed consent and baseline survey completion, participants were randomized in a 4:4:2 ratio to receive high-dose *V. atypica* (1×10¹⁰ CFU per capsule), low-dose *V. atypica* (5×10⁹ CFU per capsule), or placebo. The study employed a single-blind design in which participants were unaware of their treatment assignment, but investigators and study staff had access to allocation information.

#### Study Product

*Veillonella atypica* FB0054 capsules contained lyophilized bacteria with microcrystalline cellulose and hypromellose as excipients, encapsulated in acid-resistant hydroxypropyl methylcellulose (HPMC) capsules to enhance delivery to the colon. Placebo capsules contained microcrystalline cellulose and hypromellose in identical acid-resistant capsules, indistinguishable in appearance from active treatment. Capsules were manufactured under Good Manufacturing Practice (GMP) conditions and shipped directly to participants’ homes. Participants were instructed to take one capsule daily, preferably at the same time each day, and to store capsules at room temperature.

#### Study Procedures

The 8-week study comprised three phases (**Figure 1**): a 2-week baseline observation period (weeks 1-2), a 4-week supplementation period (weeks 3-6), and a 2-week washout period (weeks 7-8). All study activities were conducted remotely via the CHLOE smartphone application.

During the baseline period, participants completed a comprehensive health and lifestyle survey capturing demographics, medical history, exercise habits, dietary patterns, and baseline fatigue levels. Throughout all three study phases, participants completed: (1) **daily surveys** assessing sleep duration and quality (5-point Likert scale), energy level (binary: high energy day yes/no), and bowel movements (count); and (2) **weekly surveys** assessing fatigue interference (“How many days in the prior week did fatigue interfere with everyday activities?”, 0-7 scale), physical activity hours (“How many total hours did you engage in physical activity this week?”), physical activity frequency and intensity (Likert scales), mood, recovery, digestive health, and general quality of life (all Likert scales).

At the conclusion of baseline (end of week 2) and supplementation (end of week 6) periods, participants completed two validated questionnaires: the Multidimensional Fatigue Inventory (MFI-20), which assesses five dimensions of fatigue (General Fatigue, Physical Fatigue, Reduced Activity, Reduced Motivation, Mental Fatigue) with each subscale scored 4-20 and total scores ranging 20-100; and the PROMIS General Self-Efficacy 4-item short form. At study completion (end of week 8), participants completed a final survey assessing overall changes observed during the study and open-ended questions regarding study experience.

#### Adverse Event Monitoring

Adverse events (AEs) were monitored through weekly surveys asking participants whether they experienced any symptoms or stopped supplementation due to side effects, with severity and symptom details captured via free-text and structured responses. Participants could also report AEs directly to study staff by a toll-free phone number or email at any time. Serious adverse events (SAEs) were defined as events that were life-threatening, required hospitalization, resulted in disability or permanent damage, or constituted other important medical events. AE monitoring and reporting were managed by SafetyCall International, which provided 24/7 telephonic adverse event triage, documentation, and medical consultation services. No SAEs were reported.

#### Compliance and Compensation

Compliance was assessed by self-reported supplement use captured in weekly surveys. To be included in primary analyses, participants were required to complete ≥50% of surveys in each of the three study phases (baseline, supplementation, washout). Participants received in-app rewards (Chloe points redeemable for Amazon gift cards) for survey completion. Those completing ≥50% of surveys in all three phases received a 50% discount coupon for future *Veillonella* product purchases. Participants randomized to placebo who met completion criteria additionally received a coupon for one month of free *Veillonella* supplementation upon commercial product launch.

### MOUSE STUDY

#### Ethics Statement and Animal Husbandry

All animal procedures were approved by the Joslin Diabetes Center Institutional Animal Care & Use Committee (IACUC Protocol 2021-02, approved January 13, 2022) and complied with all relevant ethical regulations. Male C57BL/6J mice (4 months old at study onset; The Jackson Laboratory) were maintained on a 12-hour light-dark cycle under constant temperature (22 ± 2°C) and humidity (40–60%) with ad libitum access to water and standard rodent chow (LabDiet 5053).

#### Experimental Design and Supplementation

Mice were single-housed with continuous access to running wheels (see Voluntary Wheel Running section below). Following a ∼3-week acclimation period to establish stable baseline running behavior, animals were randomized into three groups (n=8 per group): PBS (vehicle control), *Veillonella atypica* FB0054, or *Lactobacillus johnsonii*. To avoid gavage-induced stress, supplements were delivered in peanut butter vehicle. Mice were first acclimated to peanut butter consumption (5 g per mouse of organic, no-sugar-added, no-fat peanut butter) over three consecutive days to ensure complete consumption.

Lyophilized probiotic powders were quantified by colony-forming unit (CFU) assays on BHI-lactate agar plates. Daily doses equivalent to 1×10¹⁰ CFU were prepared by thoroughly mixing probiotic powder or PBS into 5 g peanut butter per mouse. Fresh supplement-peanut butter mixtures were prepared daily during the first week and three times weekly thereafter. Supplementation continued for 50 days (Day 0–Day 49), followed by a 34-day observation/washout period (Day 50–Day 83; total post-Day 0 monitoring: 84 days). Body weight, running wheel activity, and fecal samples were monitored throughout. Fecal samples were collected weekly, snap-frozen, and stored at −80°C until DNA extraction.

#### Voluntary Wheel Running

The voluntary exercise was performed in single-housed cages with free access to a running wheel (Columbus Instruments Multi-Device Instruments). Before the experiment and data acquisition, mice were given three days of adjustment to the single-housed setup and wheel presence. Wheel revolutions were continuously monitored and recorded, and data was obtained every 10 min.

#### Dopamine ELISA

Striata were collected and snap-frozen and stored at −80 °C. Brains were homogenized in ice-cold PBS and centrifuged at 5,000g for 10 min at +4 °C, and then 50 μl of supernatant was used for quantification of dopamine in brain tissue following the manufacturer’s instructions (BioVision, K4219-100).

#### DNA stool extraction and microbiota analysis

Fresh stools were collected into 2 ml tubes, snap-frozen, and stored at −80 °C until DNA extraction. Total genomic DNA (gDNA) was isolated with ZymoBIOMICS™ DNA Miniprep Kit (Zymo Research, Irvine, CA, United States) according to the manufacturer’s instructions. Isolated DNA was checked for concentration using a Nanodrop spectrophotometer. Quantification of bacterial DNA was performed by quantitative real time PCR (q-RTPCR) analysis with the SYBR Green PCR master mix (Applied Biosystem, Foster City, CA, USA). All assays were performed using an Applied Biosystems QuantStudio 6.

### STATISTICAL ANALYSIS

All analyses were performed in R version 4.5.1. Statistical significance was defined as two-tailed *p* < 0.05 unless otherwise specified. For the human trial, the primary analytic cohort comprised participants completing ≥50% of assessments per study phase (n=151). High-dose and low-dose *V. atypica* groups were pooled after preliminary analyses confirmed similar response patterns across doses.

Longitudinal assessments of fatigue interference and physical activity hours employed linear mixed-effects models (lme4 package) adjusting for individual baseline values (calculated as mean of weeks 1-2) and including random intercepts for participants. Models were fit to post-baseline data (weeks 3-8) to estimate treatment-specific trajectories. Pre/post MFI comparisons used linear mixed models with treatment, time, and treatment×time interaction as fixed effects and random intercepts for participants. Daily sleep quality employed difference-in-differences analysis comparing baseline (days 0-13) to supplementation (days 14-41) or washout (days 42-56) periods. Adverse events were compared between groups using χ² tests.

For mouse voluntary wheel running, linear mixed-effects models included group, day (continuous post-supplementation), and group×day interaction as fixed effects, with random intercepts and slopes for each mouse. Striatal dopamine concentrations were compared by one-way ANOVA with Tukey’s post-hoc tests. Colonization was assessed by qPCR with group comparisons at Day 50 using one-way ANOVA.

No corrections for multiple comparisons were applied to the prespecified primary and secondary endpoints, as these represented distinct hypotheses rather than multiple tests of a single hypothesis. Exploratory analyses are noted as such in Results.

## Data Availability

All data produced in the present work are contained in the manuscript

## SUPPLEMENTARY FIGURES & TABLES

**Supplementary Figure 1.**
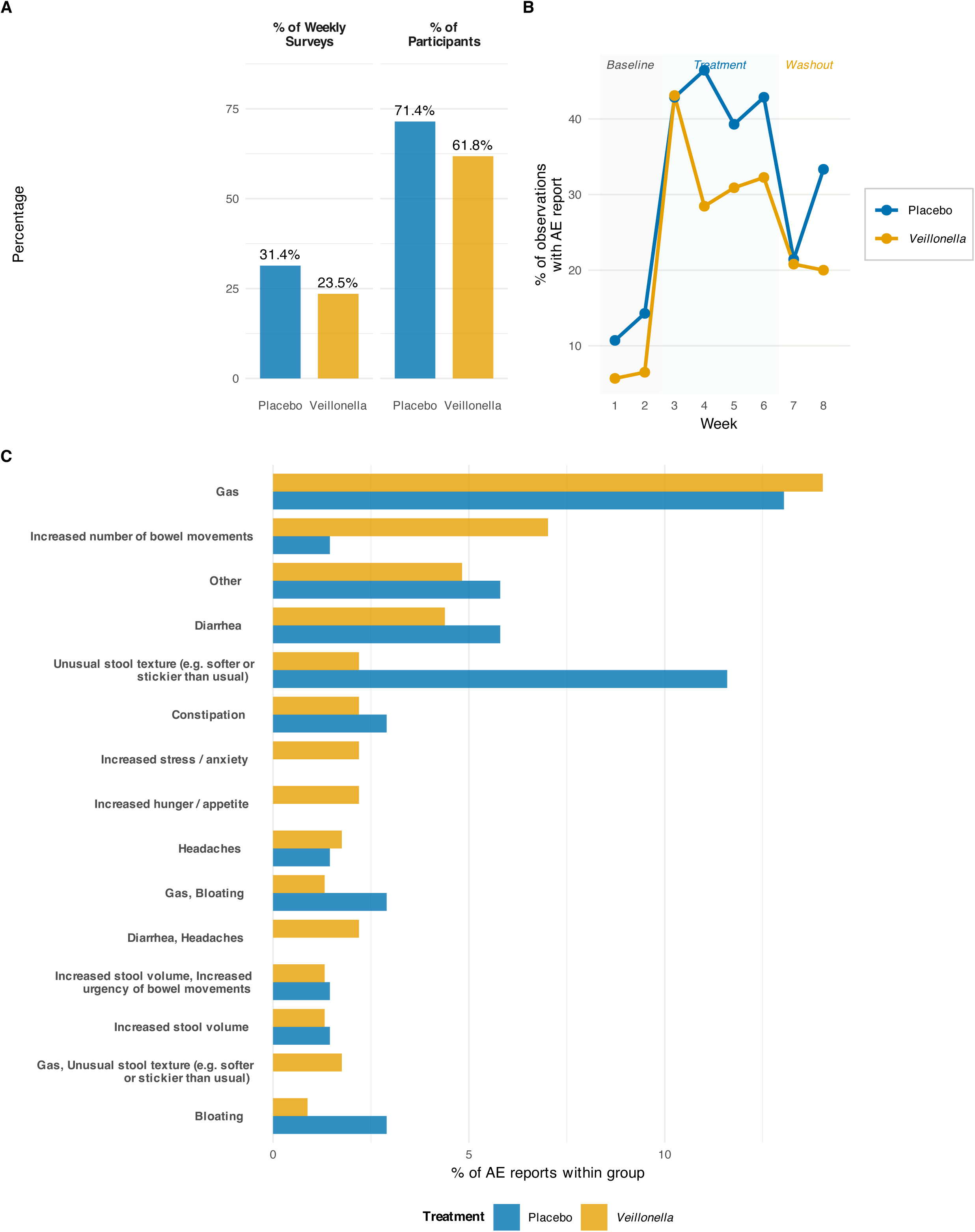
Adverse event reporting rates and patterns during *Veillonella* supplementation trial. Adverse event reporting rates and patterns during *Veillonella* supplementation trial. **(A)** Overall adverse event (AE) reporting rates by treatment group, shown as percentage of weekly survey observations reporting any AE (left) and percentage of participants who reported at least one AE during the study (right). Both groups showed similar reporting rates (Placebo: 31.4% of observations, 71.4% of participants; Veillonella: 23.5% of observations, 61.8% of participants), indicating comparable safety profiles. **(B)** Temporal pattern of AE reporting across the 8-week study period. The percentage of weekly observations with AE reports is plotted for both treatment groups across baseline (weeks 1-2, gray), supplementation (weeks 3-6, light blue), and washout (weeks 7-8, light yellow) periods. AE reporting increased during the treatment phase in both groups, peaking around weeks 3-5, then declining during washout. Both groups showed similar temporal patterns with no significant between-group differences. **(C)** The 15 most frequently reported side effects pooled across both treatment groups, stratified by treatment assignment. Bar heights represent the percentage of AE reports within each group that mentioned each specific side effect. The most common AEs were gastrointestinal in nature (gas, bloating, constipation, diarrhea), consistent with probiotic supplementation effects. The distribution of specific AE types was similar between Placebo and Veillonella groups, with gas being the most frequently reported symptom in both groups. All reported AEs were mild in severity; no serious adverse events occurred. These findings indicate that *Veillonella* supplementation was well-tolerated with a safety profile comparable to placebo.

**Supplementary Figure 2.**
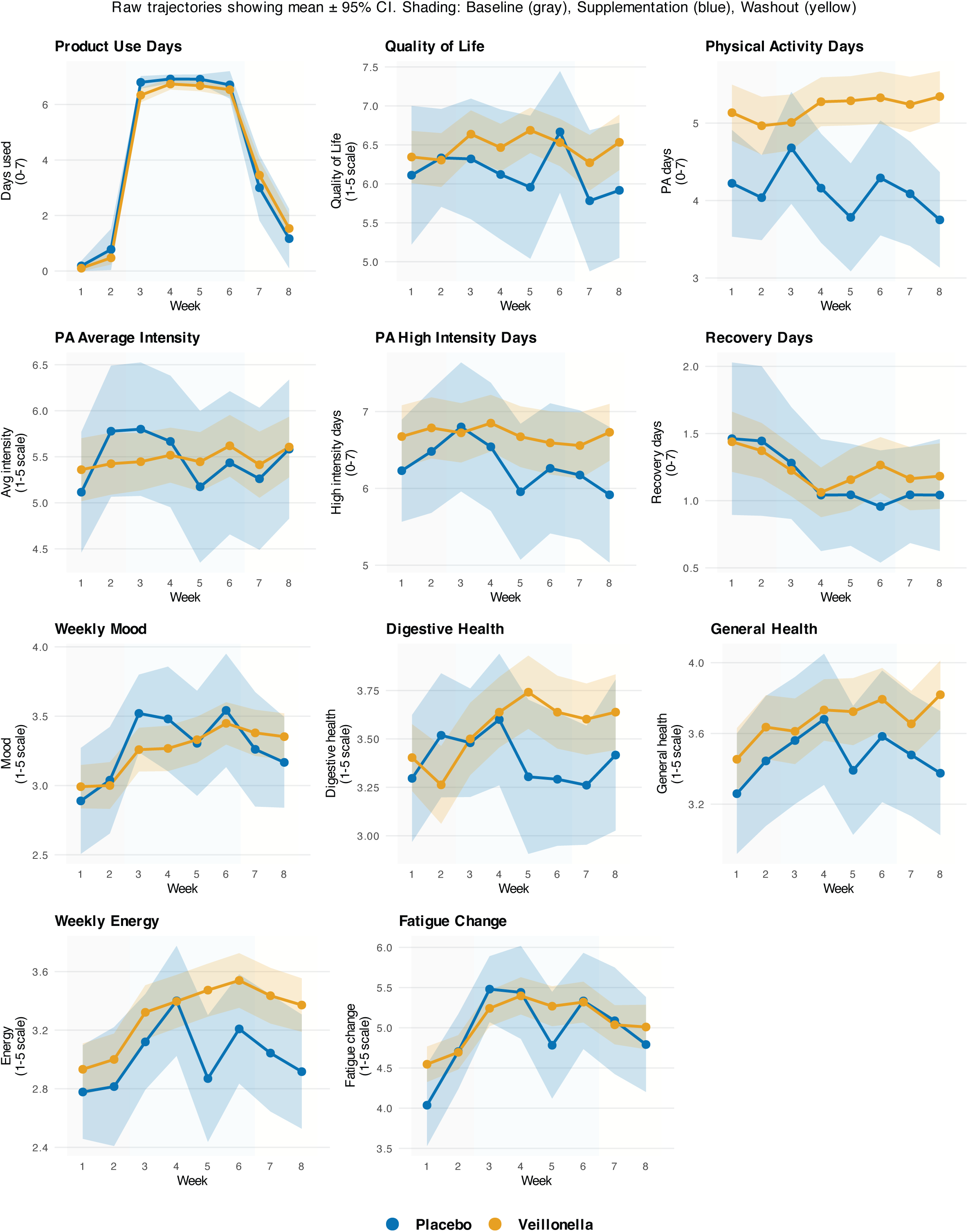
Weekly survey measures showing no treatment-specific effects. Longitudinal trajectories for additional weekly survey measures assessed throughout the 8-week study. Points represent group means (Placebo: blue; *Veillonella*: orange); error bars show 95% confidence intervals calculated from standard errors. None of these measures showed significant treatment×week interactions in linear mixed-effects models (all *p* > 0.15), indicating that treatment effects on fatigue interference and physical activity hours (Figure 2 and Figure 3) were specific rather than reflecting global wellness improvements. Measures include: **Quality of Life** (QoL; 1-5 Likert scale), **Product Use Days** (days per week supplement taken), **Physical Activity Days** (Phys A Days; days per week with any physical activity), **Physical Activity Average Intensity** (Phys A Avg Intensity; 1-10 scale), **Physical Activity Feeling** (Phys A Feeling; 1-5 scale assessing enjoyment), **Physical Activity High Intensity** (Phys A High Intensity; 1-10 scale), **Recovery Days** (days per week feeling fully recovered), **Mood Weekly** (1-5 Likert scale), **Digestive Health Weekly** (1-5 Likert scale), **General Health Weekly** (1-5 Likert scale), **Energy Weekly** (1-5 Likert scale), and **Fatigue Change Weekly** (change in fatigue relative to previous week; 1-7 scale). Vertical lines demarcate study phases. The lack of treatment differences across these diverse measures, coupled with specific effects on fatigue interference and activity hours, argues against non-specific placebo or engagement effects and supports a targeted mechanism related to exercise motivation. *Veillonella*, n = 119; Placebo, n = 27.

**Supplementary Figure 3.**
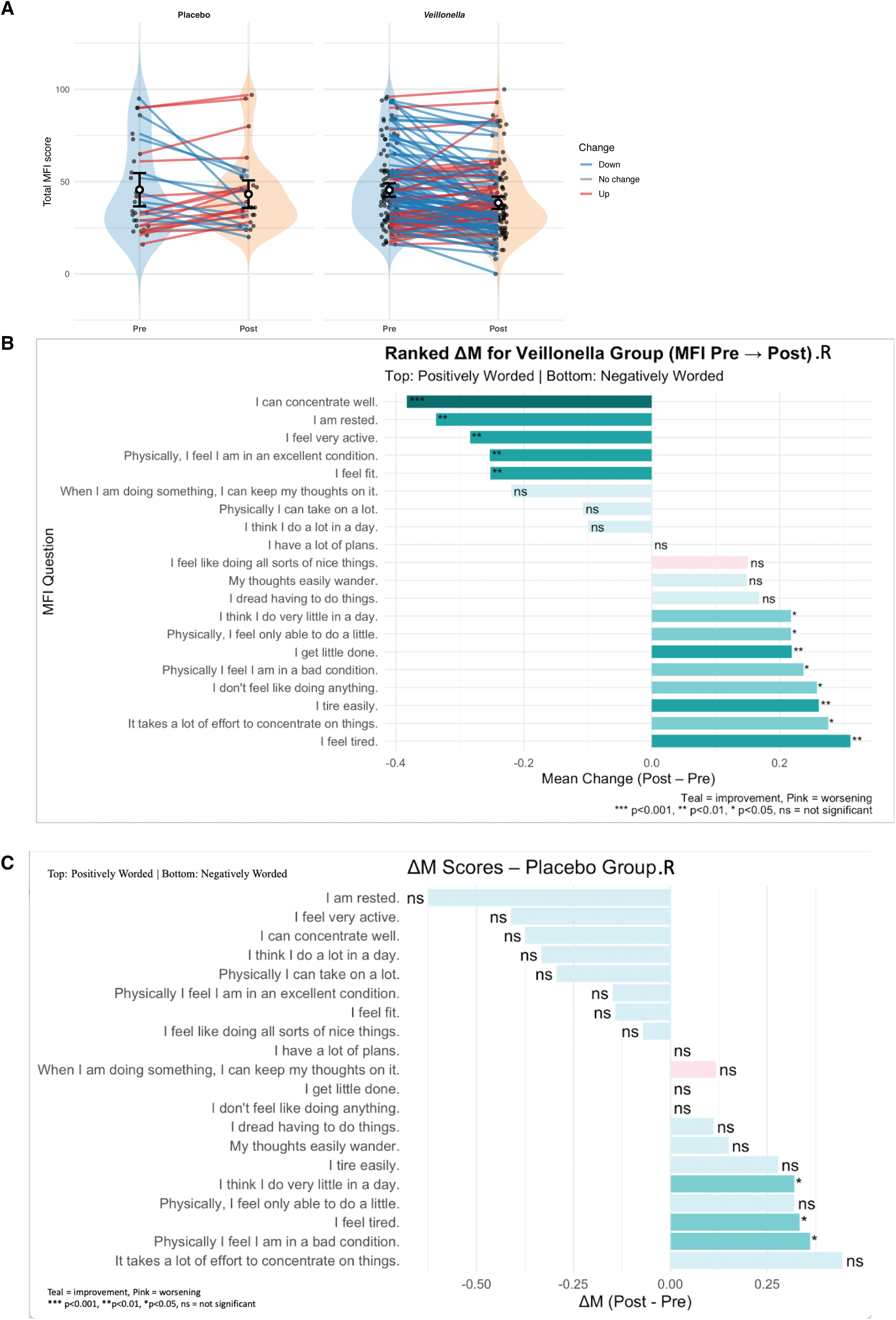
Pre- and post-supplementation MFI scores show comparable improvements in both treatment groups, with item-level analysis revealing question-specific response patterns. (**A**) Violin plots display the distribution of Total MFI scores (sum of five subscales; range 20–100) at baseline (Pre; week 0) and post-supplementation (Post; week 6) for placebo (left panel) and *Veillonella* (right panel) groups. Individual participant trajectories are shown as connected points, color-coded by direction of change: blue = decreased fatigue (improvement), gray = no change, red = increased fatigue (worsening). Open circles with error bars represent group means ± SEM. Both groups exhibited substantial reductions in Total MFI (placebo: −6.2 ± 1.6 points; *Veillonella*: −7.0 ± 1.2 points), but the treatment×time interaction was not significant (*p*=0.66), indicating parallel improvement trajectories. The majority of participants in both groups showed decreased fatigue (blue trajectories), with relatively few showing increased fatigue or no change. (**B**) Item-level analysis for the *Veillonella* group showing mean change scores (Post − Pre) for each of the 20 MFI questions, ranked from most positive (top) to most negative (bottom). Questions are ordered to group positively-worded items (top section) and negatively-worded items (bottom section). Teal bars indicate improvement (negative change = reduced fatigue); pink bars indicate worsening (positive change = increased fatigue). Within-group paired t-tests: \*\*\**p*<0.001, \*\**p*<0.01, \**p*<0.05, ns = not significant. Most items showed significant improvement, with the largest effects for “I feel tired” and “It takes a lot of effort to concentrate on things.” (**C**) Item-level analysis for the placebo group, displayed in identical format to panel B. The placebo group also showed significant improvements on multiple items, with similar questions showing the largest effects. The comparable patterns across panels B and C support the conclusion that both groups improved in parallel, consistent with the non-significant treatment×time interaction for total scores. Linear mixed-effects model: Total_MFI ∼ treatment × time + (1|participant_id). Placebo, n=28; *Veillonella*, n=123.

**Supplementary Figure 4.**
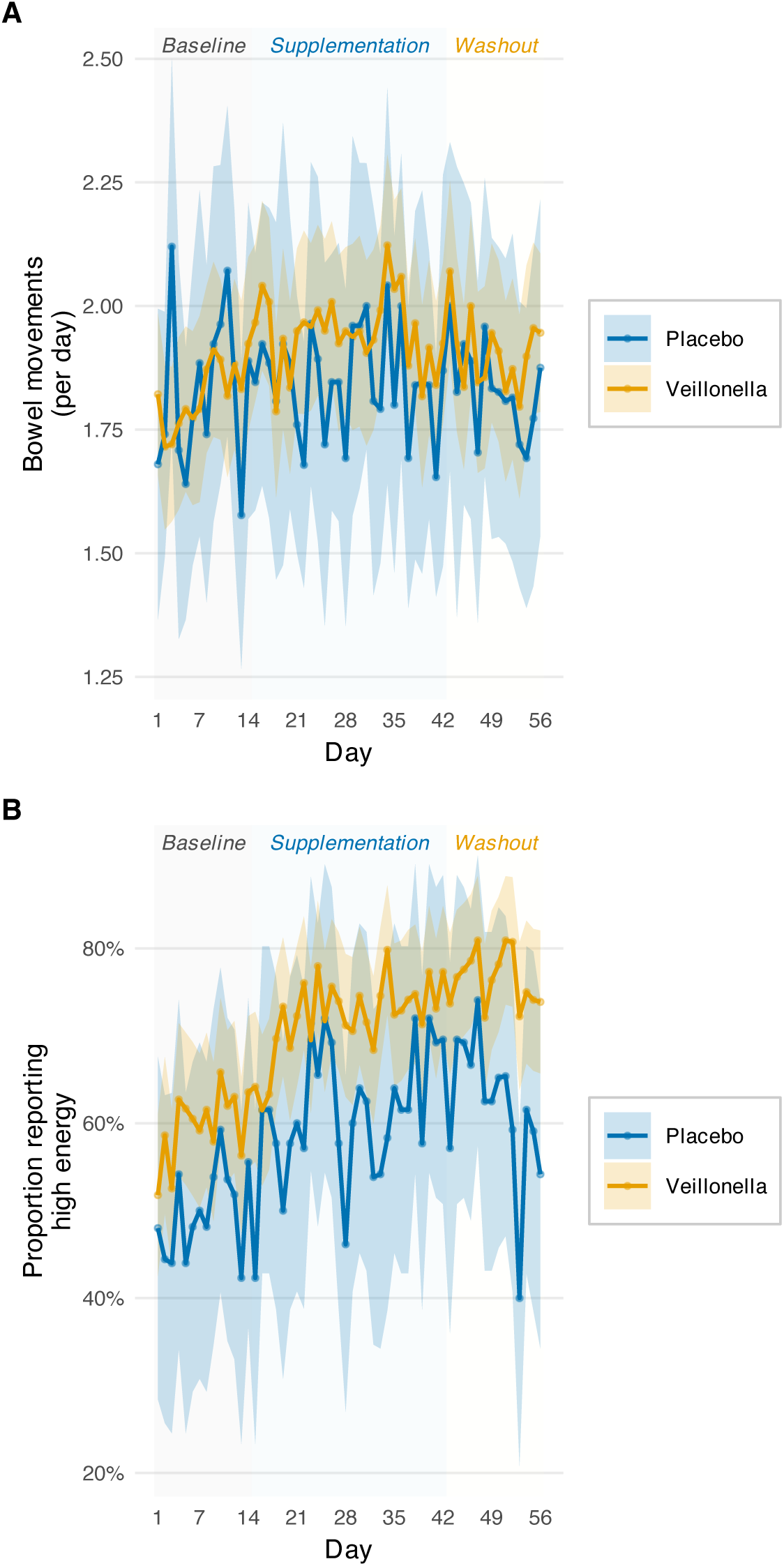
Daily measures of bowel movements and energy levels. (**A**) Daily bowel movements (count per day) across the study period (days 0–56) for placebo (blue) and *V. atypica* (orange) groups. Individual daily observations shown as semi-transparent points; smoothed group trajectories (LOESS) shown as bold lines with 95% confidence bands. No treatment×period interactions were detected for either supplementation (*p*=0.88) or washout (*p*=0.72) periods. Mean bowel movements: placebo 1.93±0.08/day; *V. atypica* 1.98±0.05/day. The absence of gastrointestinal effects is consistent with adverse event reporting (Supplementary Figure 1) and supports the tolerability of *V. atypica* supplementation. (**B**) Proportion of participants reporting “high energy” days (binary outcome) across the study. Lines show smoothed group proportions (LOESS) with 95% confidence bands. The *V. atypica* group showed consistently higher proportions of high-energy days throughout the study, with trends most pronounced during supplementation and washout. Mixed-effects logistic regression revealed a trend during washout (OR=2.36; 95% CI 0.93–6.02; *p*=0.071) that did not reach statistical significance. During supplementation: OR=1.48 (*p*=0.27). Baseline proportions were similar (placebo: 62%; *V. atypica*: 65%; *p*=0.58). The marginal statistical significance and lack of effect during active supplementation warrant cautious interpretation. Vertical shaded regions demarcate study phases: Baseline (gray, days 0–13), Supplementation (blue, days 14–41), and Washout (yellow, days 42–56). *V. atypica*, n=119; Placebo, n=27.

**Supplementary Table 1.** Multidimensional Fatigue Inventory (MFI-20) subscale and total scores at baseline and post-supplementation. Each row presents one of the five MFI-20 subscales (General Fatigue, Physical Fatigue, Reduced Activity, Reduced Motivation, Mental Fatigue) or the Total MFI score (sum of subscales). Columns show baseline mean ± SEM, post-supplementation mean ± SEM, and within-group change (post − baseline) for placebo and *Veillonella* groups separately. The final column reports the treatment×time interaction *p*-value from linear mixed-effects models (formula: outcome ∼ treatment × time + (1|participant_id)), testing whether the rate of change differed between groups. All subscales and Total MFI showed significant within-subject improvements over time (time main effects: all *p*<0.01), but no treatment×time interactions reached significance (all *p*>0.40), indicating comparable improvement trajectories. Subscale scores range from 4 (low fatigue) to 20 (high fatigue); Total MFI ranges from 20 to 100. Placebo, n=28; *Veillonella*, n=123.

| Summary of MFI Results: Veillonella RCT |  |  |  |  |  |  |  |  |  |  |
| --- | --- | --- | --- | --- | --- | --- | --- | --- | --- | --- |
| Multidimensional Fatigue Inventory scores at baseline (Pre) and week 4 (Post) |  |  |  |  |  |  |  |  |  |  |
| MFI Domain | Placebo |  |  |  | Veillonella |  |  |  | Treatment Effect |  |
|  | N | Pre | Post | Change | N | Pre | Post | Change | Hedges g (95% CI) | P-value* |
| General Fatigue (subscale) | 21 | 14.1 ± 4.4 | 12.2 ± 4.6 | -1.9 ± 3.6 | 97 | 12.4 ± 4.3 | 10.6 ± 4.9 | -1.5 ± 2.9 | -0.11 (-0.59, 0.36) | 0.625 |
| Physical Fatigue (subscale) | 22 | 10.9 ± 5.4 | 9.3 ± 4.8 | -1.3 ± 3.0 | 94 | 9.8 ± 5.2 | 8.6 ± 4.9 | -1.2 ± 2.4 | -0.04 (-0.50, 0.43) | 0.828 |
| Reduced Activity (subscale) | 17 | 13.7 ± 5.4 | 11.9 ± 5.0 | -0.6 ± 2.7 | 80 | 12.1 ± 5.1 | 10.7 ± 4.8 | -1.0 ± 3.0 | 0.11 (-0.42, 0.64) | 0.635 |
| Reduced Motivation (subscale) | 21 | 10.7 ± 5.3 | 9.2 ± 4.7 | -0.5 ± 2.2 | 99 | 9.5 ± 4.8 | 8.3 ± 4.4 | -0.8 ± 2.2 | 0.15 (-0.33, 0.62) | 0.508 |
| Mental Fatigue (subscale) | 15 | 10.1 ± 4.2 | 8.8 ± 4.0 | 0.3 ± 2.7 | 81 | 9.5 ± 4.1 | 8.5 ± 3.8 | -0.5 ± 2.6 | 0.29 (-0.26, 0.85) | 0.265 |
| Total MFI Score | 28 | 45.6 ± 24.3 | 43.2 ± 20.1 | -2.4 ± 17.7 | 123 | 45.4 ± 20.6 | 38.5 ± 19.0 | -6.9 ± 15.8 | 0.28 (-0.14, 0.69) | 0.156 |
| Item 1 (General: feel fit) | 28 | 3.1 ± 1.2 | 3.0 ± 1.3 | -0.1 ± 1.2 | 123 | 2.7 ± 1.3 | 2.5 ± 1.2 | -0.3 ± 1.0 | 0.14 (-0.27, 0.55) | 0.485 |
| Item 2 (Physical: physically feel) | 25 | 3.5 ± 1.3 | 4.1 ± 1.3 | 0.3 ± 0.9 | 110 | 3.8 ± 1.4 | 4.1 ± 1.3 | 0.2 ± 1.1 | 0.08 (-0.36, 0.52) | 0.559 |
| Item 3 (Motivation: feel active) | 17 | 3.2 ± 1.4 | 2.8 ± 1.3 | -0.4 ± 0.9 | 95 | 2.7 ± 1.4 | 2.3 ± 1.4 | -0.3 ± 0.9 | -0.07 (-0.59, 0.45) | 0.776 |
| Item 4 (Mental: concentrate well) | 14 | 2.8 ± 1.5 | 2.4 ± 1.4 | -0.1 ± 0.7 | 73 | 2.3 ± 1.3 | 2.2 ± 1.2 | 0.1 ± 1.0 | -0.14 (-0.71, 0.44) | 0.567 |
| Item 5 (General: feel tired) | 18 | 2.3 ± 1.4 | 2.7 ± 1.3 | 0.3 ± 0.6 | 87 | 2.4 ± 1.3 | 2.8 ± 1.4 | 0.3 ± 1.0 | 0.02 (-0.49, 0.53) | 0.898 |
| Item 6 (Motivation: do a lot) | 15 | 3.1 ± 1.4 | 2.5 ± 1.4 | -0.3 ± 0.7 | 91 | 2.5 ± 1.4 | 2.2 ± 1.3 | -0.1 ± 1.0 | -0.22 (-0.77, 0.33) | 0.416 |
| Item 7 (Activity: think about other) | 17 | 3.1 ± 1.5 | 3.1 ± 1.2 | 0.1 ± 0.8 | 82 | 2.8 ± 1.3 | 2.5 ± 1.3 | -0.2 ± 1.2 | 0.30 (-0.23, 0.83) | 0.214 |
| Item 8 (Physical: physically able) | 17 | 3.1 ± 1.5 | 2.6 ± 1.3 | -0.3 ± 1.8 | 93 | 2.4 ± 1.4 | 2.2 ± 1.3 | -0.2 ± 1.0 | -0.12 (-0.64, 0.39) | 0.600 |
| Item 9 (Mental: thinking requires effort) | 18 | 3.0 ± 1.5 | 3.5 ± 1.2 | 0.1 ± 1.2 | 83 | 3.3 ± 1.4 | 3.6 ± 1.3 | 0.2 ± 1.1 | -0.09 (-0.61, 0.42) | 0.771 |
| Item 10 (Motivation: feel like doing) | 25 | 3.6 ± 1.6 | 3.8 ± 1.3 | 0.3 ± 0.7 | 101 | 3.8 ± 1.3 | 4.2 ± 1.1 | 0.3 ± 0.9 | 0.03 (-0.41, 0.47) | 0.982 |
| Item 11 (Activity: concentrate on things) | 16 | 3.3 ± 1.4 | 2.8 ± 1.4 | -0.4 ± 1.1 | 81 | 2.9 ± 1.3 | 2.4 ± 1.2 | -0.4 ± 0.9 | 0.01 (-0.53, 0.55) | 0.809 |
| Item 12 (General: rested) | 16 | 4.0 ± 1.0 | 3.2 ± 1.3 | -0.6 ± 1.5 | 80 | 3.4 ± 1.3 | 2.9 ± 1.4 | -0.3 ± 1.0 | -0.25 (-0.80, 0.29) | 0.359 |
| Item 13 (Activity: concentrate to do) | 18 | 2.9 ± 1.5 | 3.5 ± 1.3 | 0.4 ± 0.9 | 87 | 3.0 ± 1.5 | 3.4 ± 1.3 | 0.3 ± 1.2 | 0.15 (-0.36, 0.66) | 0.485 |
| *P-value from treatment × time interaction in mixed-effects model |  |  |  |  |  |  |  |  |  |  |
| Values shown as mean ± SD. Change = Post – Pre. |  |  |  |  |  |  |  |  |  |  |
| Hedges g: effect size for between-group difference in change scores. |  |  |  |  |  |  |  |  |  |  |

Summary of MFI Results: Veillonella RCT Multidimensional Fatigue Inventory scores at baseline (Pre) and week 4 (Post)
| MFI Domain | Placebo |  |  |  | Veillonella |  |  |  | Treatment Effect |  |
| --- | --- | --- | --- | --- | --- | --- | --- | --- | --- | --- |
|  | N | Pre | Post | Change | N | Pre | Post | Change | Hedges g (95% CI) | P-value* |
| Item 14 (Physical: condition is good) | 25 | 3.5 ± 1.5 | 3.8 ± 1.3 | 0.4 ± 0.7 | 97 | 3.8 ± 1.4 | 4.1 ± 1.3 | 0.2 ± 0.9 | 0.14 (-0.30, 0.58) | 0.554 |
| Item 15 (Mental: thoughts wander) | 20 | 1.9 ± 1.1 | 1.9 ± 1.1 | 0.0 ± 1.3 | 92 | 2.1 ± 1.2 | 2.0 ± 1.2 | -0.0 ± 0.9 | 0.02 (-0.46, 0.51) | 0.888 |
| Item 16 (General: get a lot done) | 18 | 2.8 ± 1.6 | 3.3 ± 1.4 | 0.3 ± 0.7 | 88 | 3.2 ± 1.4 | 3.7 ± 1.4 | 0.4 ± 0.8 | -0.12 (-0.63, 0.39) | 0.590 |
| Item 17 (Motivation: can concentrate) | 25 | 3.8 ± 1.2 | 3.8 ± 1.4 | 0.0 ± 1.0 | 96 | 3.8 ± 1.3 | 4.0 ± 1.2 | 0.2 ± 0.7 | -0.29 (-0.73, 0.15) | 0.175 |
| Item 18 (Mental: feel relaxed) | 23 | 3.6 ± 1.3 | 3.7 ± 1.4 | 0.0 ± 1.0 | 93 | 3.5 ± 1.4 | 3.9 ± 1.3 | 0.3 ± 1.0 | -0.29 (-0.75, 0.17) | 0.180 |
| Item 19 (Activity: takes effort) | 20 | 2.6 ± 1.2 | 2.7 ± 1.4 | 0.2 ± 1.0 | 81 | 2.6 ± 1.5 | 2.9 ± 1.4 | 0.2 ± 0.9 | -0.03 (-0.52, 0.47) | 0.888 |
| Item 20 (Physical: do very little) | 20 | 3.1 ± 1.5 | 2.9 ± 1.3 | -0.2 ± 0.7 | 87 | 2.9 ± 1.5 | 2.7 ± 1.5 | -0.3 ± 0.9 | 0.12 (-0.37, 0.61) | 0.561 |

\*P-value from treatment × time interaction in mixed-effects model
Values shown as mean ± SD. Change = Post – Pre.
Hedges g: effect size for between-group difference in change scores.

## Notes

### Competing Interest Statement

A.D.K., J.R.S., and C.B. are co-founders of and hold equity in FitBiomics, Inc. M.S. was an employee of FitBiomics, Inc. P.O., N.C., and M.C.E. are employees of and hold equity in PeopleScience, Inc. E.A., N.C., K.W., and L.D.P. declare no competing interests.

### Clinical Trial

NCT06141343

### Clinical Protocols

https://clinicaltrials.gov/expert-search?term=NCT06141343

### Funding Statement

This work was funded by FitBiomics Inc. (human clinical trial), an NIH R01-DK129850 (PI: A.D.K and Sarah Lessard), and an NIH/NIDDK Diabetes Research Center grant (PI: Jean E. Schaffer).

### Author Declarations

Advarra Institutional Review Board

### Summary of Updates

Author name added, and the affiliations are fixed.

